# Dietary Intake Mendelian Randomization: Assessment and Development of Methods for Instrument Selection and Robust Inference

**DOI:** 10.1101/2025.06.26.25330002

**Authors:** Kristen J. Sutton, Julie Gervis, Moomal Jatoi, Liang-Dar Hwang, Audrey Hendricks, Debashis Ghosh, Kenneth Westerman, Joanne B. Cole

**Affiliations:** Department of Biomedical Informatics, University of Colorado Anschutz Medical Campus, Aurora, Colorado, 80045, USA; Center for Genomic Medicine, Massachusetts General Hospital, Boston, MA; Institute for Molecular Bioscience, University of Queensland, Brisbane, Queensland, 4072, Australia; Department of Mathematical and Statistical Sciences, University of Colorado Denver, Denver, Colorado, 80217, USA; Department of Biostatistics & Informatics, University of Colorado Anschutz Medical Campus, Aurora, Colorado, 80045, USA; Department of Medicine, Clinical and Translational Epidemiology Unit, Mongan Institute, Massachusetts General Hospital, Boston, MA, USA; Metabolism Program, Broad Institute of MIT and Harvard, Cambridge, MA, USA; Department of Medicine, Harvard Medical School, Boston, MA, USA

## Abstract

**Background:** Mendelian randomization (MR) uses genetic instruments (GI) to infer causality between exposures, like dietary intake, and health outcomes. Almost all MR of dietary intake use the full set of genome-wide significant (GWS) variants in the GI, and therefore, causal estimates are likely biased by variants that act indirectly on diet.

**Objective:** First, we performed an assessment of the diet MR literature to evaluate the applications and approaches common in the field. Second, using conventional two-sample MR techniques with GWS variants, we evaluated whether MR could detect expected associations between six diet-health relationships supported by existing nutrition science literature. Third, we developed and tested methods for refining the GI using filtering and mediation-based approaches.

**Methods:** Studies that performed MR of foods or beverages on any health outcome were identified in PubMed. We recorded how the GI was created, what dietary intake traits were studied, how the exclusion restriction assumption was evaluated, and what sensitivity tests were performed. We tested if conventional MR methods could detect established diet-health relationships by selecting a biomarker and disease outcome for each dietary trait (six positive controls total). This included oily fish intake on triglycerides (TG) and cardiovascular disease (CVD), alcohol intake on alanine aminotransferase (ALT) and liver cirrhosis, and white vs whole grain or brown bread on LDL cholesterol (LDL-C) and CVD. To refine the GI to better estimate the direct effect of diet by removing or accounting for the indirect effects of confounders, we tested two phenome-wide association study (PheWAS) based GI filtering approaches and a mediation approach via multivariable MR (MVMR). Causal inferences were estimated by the inverse variance weighted (IVW) and weighted median (WM) estimators and by MR-CAUSE.

**Results:** There is a strong and rapidly expanding interest in applying MR to dietary intake exposures (178 studies identified with 76 published in 2024). Existing studies showed a wide range of methodological rigor, especially with respect to GI specificity, which raised concerns whether MR using GWS GIs can adequately evaluate diet-health relationships. In empirical testing, conventional two-sample MR methods on GWS GIs only identified the relationships between oily fish on TG and white vs whole grain or brown bread on LDL-C using the WM estimator, whereas no relationships were identified by the IVW estimator. Filtering the GI improved the ability to detect the expectation for diet-biomarker pairs (IVW, oily fish on TG: ß=-0.12 [95% CI –0.18 to –0.054]; IVW, white vs whole grain or brown bread on LDL-C: ß = 0.11 [95% CI 0.058 to 0.16]) but not diet-disease pairs. MR-CAUSE identified the only diet-disease association – white vs. whole grain or brown bread on CVD (γ=0.17 [95% credible interval, 0.09 to 0.25]). Furthermore, MR-CAUSE found that many diet-health relationships were impacted by confounding. We evaluated which traits contributed to confounding via the PheWAS results and found that body composition traits were the most prevalent confounders. The PheWAS output was used to prioritize traits for MVMR and rescued the expected direct effect of alcohol on ALT (ß= 0.028 [95% CI 0.017 to 0.039]).

**Conclusion:** MR studies of diet’s causal role in health have flooded the literature; however, our inconsistent associations with positive and negative controls using multiple tests and filtering methods signal a need for caution. More thoughtful curation of the GI is critical to reduce confounding due to health and environmental factors when evaluating the causal effect of diet on health.

## Introduction

Dietary intake has profound effects on health. However, many relationships between foods and health outcomes remain controversial, and multiple robust associations from observational studies have failed in randomized controlled trials [1, 2]. Mendelian randomization (MR) is a form of instrumental variable analysis that has been likened to a randomized controlled trial by leveraging the random assignment of alleles at meiosis to estimate causal effects of an exposure, like dietary intake, on health outcomes. MR promises to be a cost-effective intermediate tool to identify and clarify how foods affect health prior to undertaking long and expensive randomized trials.

In order for MR to accurately estimate causal effects, three main conditions must be satisfied: 1) relevance: the GI is associated with the exposure; 2) exchangeability: there are no confounders on the exposure-outcome relationship; and 3) exclusion restriction: there are no paths from the GI to the outcome that do not go through the exposure [3].

In recent years, multiple genome-wide association studies (GWAS) of food liking [4] and dietary intake [5–7] have identified hundreds of genetic associations. However, many of the loci associated with diet also show associations with socioeconomic status, physical activity levels, and health, raising the question of whether these genetic loci act directly on dietary intake or indirectly through other traits. As these factors independently affect health outcomes, their overlap at diet-associated loci may lead to violating the exclusion restriction assumption underlying MR and biasing the causal estimates.

This recent availability of GWAS on a variety of diet-related traits and the ease of performing two-sample MR analyses with user-friendly R packages and an online portal (MRbase.org) have led to a surge of diet MR studies – often at the expense of careful consideration of the exclusion restriction assumption. Therefore, we critically assessed the diet MR literature to characterize and assess the rigor of current approaches. Motivated by the findings of the literature assessment, we aimed to establish whether conventional two-sample MR methods using GIs composed of the full set of genome-wide significant (GWS; *P*<5×10^−8^) loci could detect established and experimentally supported diet-health relationships. Finally, we tested a suite of methods designed to be robust to pleiotropy to isolate the direct effect of diet on health independent of the indirect effects of confounders. This included MR-CAUSE, various GI filtering approaches, and multivariable MR (MVMR). Our approaches utilized publicly accessible summary-level data to test the credibility of existing GWS GIs and to provide guidance for researchers interested in applying MR to dietary intake exposures.

## Methods

An overview of the study design is presented in **Figure 1**. The STROBE-MR checklist is provided as an additional file.

**Figure 1.**
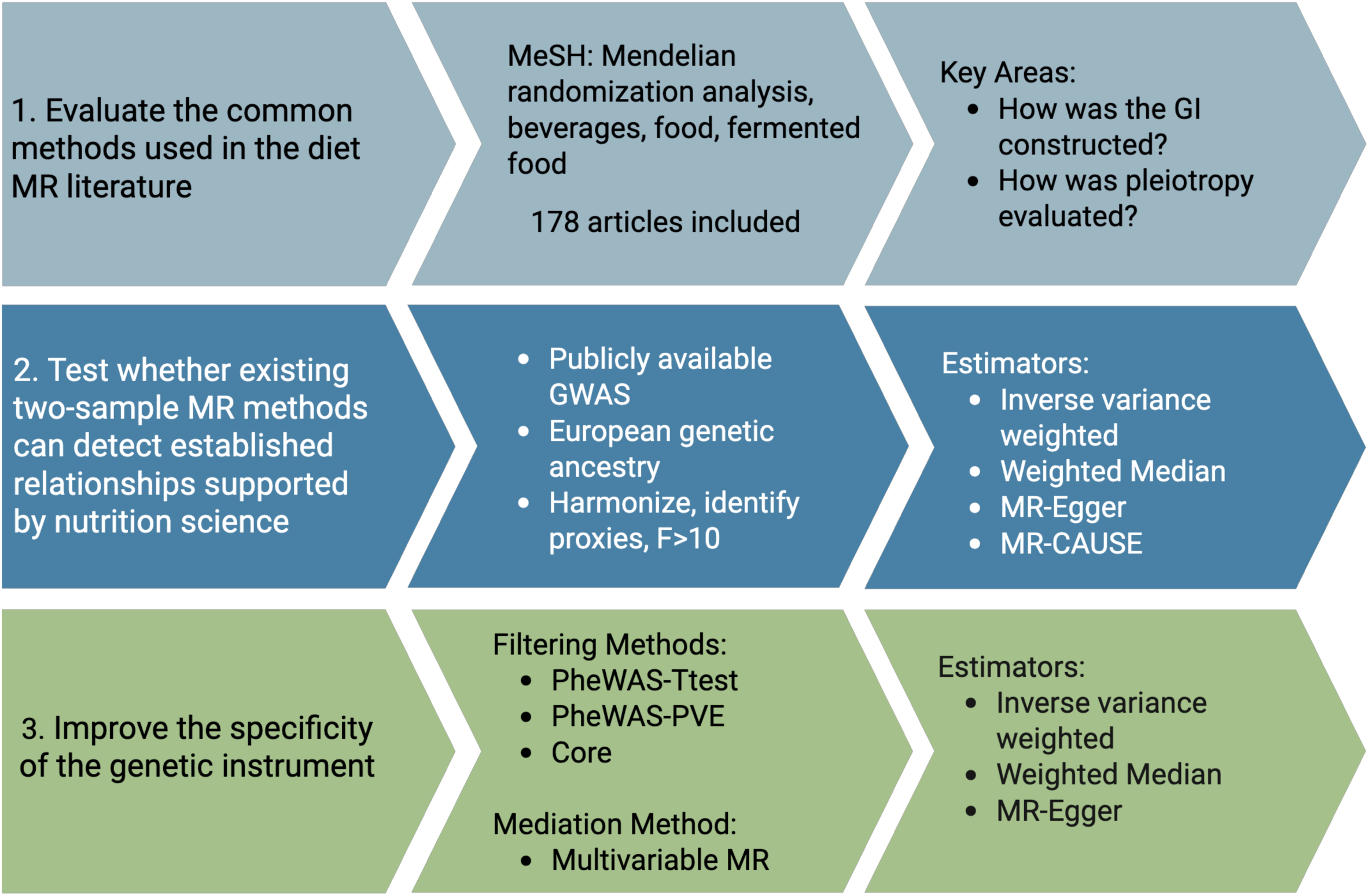
Overview of the study design.

### Assessment of the diet MR literature

The Medical Subject Headings (MeSH) terms for “Diet, Food, and Nutrition” and “Mendelian Randomization Analysis” were identified. Within the “Diet, Food, and Nutrition” hierarchy, we focused on “Food,” “Fermented Food,” and “Beverages” due to our interest in assessing the individual components of diet that affect health. The following search was completed on PubMed, “Mendelian Randomization Analysis”[Mesh] AND (“Beverages”[Mesh] OR “Fermented Foods”[Mesh] OR “Food”[Mesh]), which returned 378 articles (search date, January 24, 2025). Abstracts were reviewed by two independent researchers to identify manuscripts evaluating the relationship between a food, food group, or beverage on any health outcome. Reviews, comments, and manuscripts evaluating nutrients were removed, leaving 178 manuscripts that were read in full (**Figure S1**).

### Data extraction and synthesis

We extracted information on how the GI was constructed, how the exclusion restriction assumption was evaluated and what MR estimates and sensitivity tests were conducted. To evaluate GI composition, we recorded whether the GI consisted of variants with known functional mechanisms related to the diet trait versus variants of unknown mechanism that surpassed a statistical threshold from a GWAS. We also recorded the genetic similarity (i.e., ancestry) that the GI was built from, and whether sex-specific genetic effects were considered.

It is not possible to test the exclusion restriction assumption directly, but there are several methods that estimate or address whether the assumption is met, which we recorded. These include approaches that leverage prior knowledge, such as filtering variants from the GI if they are associated with confounding traits (i.e., PheWAS-based methods), methods that remove variants if they are statistical outliers (i.e., MR-PRESSO [8]), and methods that control for the indirect effect of confounding (i.e., MVMR [9]).

Conventional MR methods, including the estimators and sensitivity tests, were recorded. The exposures were grouped into food groups and studies that evaluated greater than five food groups were classified generally as “diet.”

### Selection of diet and health outcomes

We selected six positive control relationships between foods or beverages and health outcomes that have established nutrition science support, including plausible biological mechanisms for the effects. The prioritized foods or beverages contained a predominant nutrient of action: omega-3 fatty acids in oily fish, fiber in whole grain and brown bread, and ethanol in alcohol. To evaluate MR’s ability to detect causal relationships at different stages of complexity, we tested a biomarker and a disease outcome for each dietary exposure. We selected oily fish on triglycerides (TG) [10] and cardiovascular disease (CVD) [10], alcohol intake on alanine aminotransferase (ALT) [11, 12] and liver cirrhosis [13, 14], and bread type (white versus whole grain or brown bread) on LDL cholesterol (LDL-C) [15, 16] and CVD [17]. We used height in populations with adequate nutrition as a negative control.

### Genetic instrument selection

GWAS of oily fish, overall alcohol intake, and white vs whole grain or brown bread in 455K individuals of European genetic ancestry from the UK Biobank (UKB), adjusted for age, sex, and inverse normal transformed [5] were used to derive GIs, created from independent GWS variants (r^2^<0.8, clumping window 500 KB). Biomarker (LDL-C, ALT, TG) and disease (CVD, cirrhosis) GIs were derived from European-ancestry GWAS that maximized sample size and avoided sample overlap with the UKB [18–21]. Height GWAS summary statistics were from Loic et al., 2018 [22], as the more recent publicly available GWAS on height contained sparse genome-wide information. The selected GWAS are described in **Supplemental Table 1**, and the GIs for each dietary trait are presented in **Supplemental Table 2**. We harmonized exposure and outcome GWAS data using custom scripts to align the effect alleles between studies, remove ambiguous variants with minor allele frequency between 0.45 to 0.55, and find proxies using LDLinkR and the Great Britain reference population in build 37 (r^2^>0.8). When proxies were needed, the dietary intake variant was also replaced to avoid problems of phasing [23].

### Conventional MR tests

The inverse variance weighted (IVW) estimator with random effects and weighted median (WM) estimator were used. For the IVW tests, instrument strength was approximated using the F-statistic and heterogeneity with Cochran’s Q statistic. We used the MR-Egger intercept test to assess pleiotropy [24]. Steiger filtering was used to remove variants from the GI that explained more variance in the outcome than the exposure [25]. Following a conservative Bonferroni correction, significance was set at α=0.005 (0.05/9 trait pairs).

### Robust MR methods addressing correlated pleiotropy and imprecise genetic instruments

We applied multiple methods designed to address pleiotropy. First, we tested MR-CAUSE, a recent method that incorporates genome-wide data to test if the relationship between the exposure and outcome is causal and/or shared with a confounder (i.e., uncorrelated pleiotropy) (**Figure S2**) [26]. The MR-CAUSE nuisance parameters were calculated using 1,000,000 variants without replacement. The genome-wide variants were pruned using the ld_prune function with the 1000 Genome CEU population and a statistical threshold of 5×10^−3^.

Second, we tested two phenome-wide association study (PheWAS) GI filtering methods, which improve GI precision by identifying and removing variants with stronger effects on confounding traits from the PheWAS than dietary intake. The first method, PheWAS Two-Sample T-Test (PheWAS-Ttest), uses a two-sample t-test to remove variants if the mean effect of the variant-PheWAS-trait relationship is statistically stronger than the mean effect of the variant-diet relationship [27].

We propose and test herein a second simpler PheWAS approach, based on the same underlying comparison used by Steiger filtering. Specifically, we removed variants that explain more variance (via percent variance explained [PVE]) in any trait in the PheWAS compared to the PVE in the dietary exposure of interest. PVE was calculated using GWAS summary statistics using the following formula by Shim et al (**equation 1**) [28].

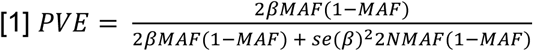

### Obtaining the PheWAS data

PheWAS data were obtained from the second round of the Neale Lab UK Biobank GWAS release, including traits related to anthropometrics, biomarkers, disease status, and lifestyle [29]. Binary traits with an effective sample size of 50,000 or inverse rank transformed quantitative traits were included (829 traits total), while traits with a genetic correlation greater than 0.75 with the dietary exposure were removed. Because dietary intake is compositional and substitutional, we performed a sensitivity PheWAS analysis in which we removed all dietary intake traits, leaving 714 traits.

### Systematically identifying traits for multivariable Mendelian randomization

The PheWAS-Ttest method generates a list of variants and their PheWAS-trait with stronger associations than with the dietary exposure of interest, suggesting their impact on diet is indirect. We developed a pipeline that uses summary-level data, genetic correlation analysis, and hierarchical clustering to systematically select representative traits from this list for inclusion in MVMR mediation analysis to tease apart diet’s direct and indirect paths on relevant health outcomes (**Figure S3**). For all confounding traits identified from the PheWAS-Ttest for each dietary exposure, pair-wise genetic correlations were calculated using LD Score Regression on LDSC formatted summary statistics from the Neale Lab [30]. Hierarchical clustering from the Euclidean distance matrix clustered these phenotypes into groups. The number of groups was determined by visually inspecting the dendrogram for biological interpretability and by evaluating the changes in branch height. For each cluster, a representative trait was selected by identifying the highest degree of variant overlap with diet. We identified GWS variants for these representative traits by clumping independent signals (r^2^=0.01, 10000 kb window) using Plink (v1.9) and the 1000 Genome Project Phase 3 European-Ancestry reference panel. MVMR models were then fit with GWS GIs including the representative traits and the original dietary exposure simultaneously.

### Software

Analyses were performed using RStudio (v 4.2.2). The libraries MendelianRandomization (v 0.10.0) [31], MR-CAUSE (v 1.2.0) [26], and MVMR (v 0.4) [9] were used to perform MR. Steiger filtering was performed using TwoSampleMR (v 0.6.16) [32].

## Results

### Assessment of the dietary intake MR literature

Our comprehensive literature search identified 178 studies since 2002 that used MR to test the causal effect of foods or beverages (and not nutrients) on health outcomes (**Supplemental Table 3**), with large increases each year over the last five years and 76 studies (∼43%) published in 2024 (**Figure 2A**).

**Figure 2.**
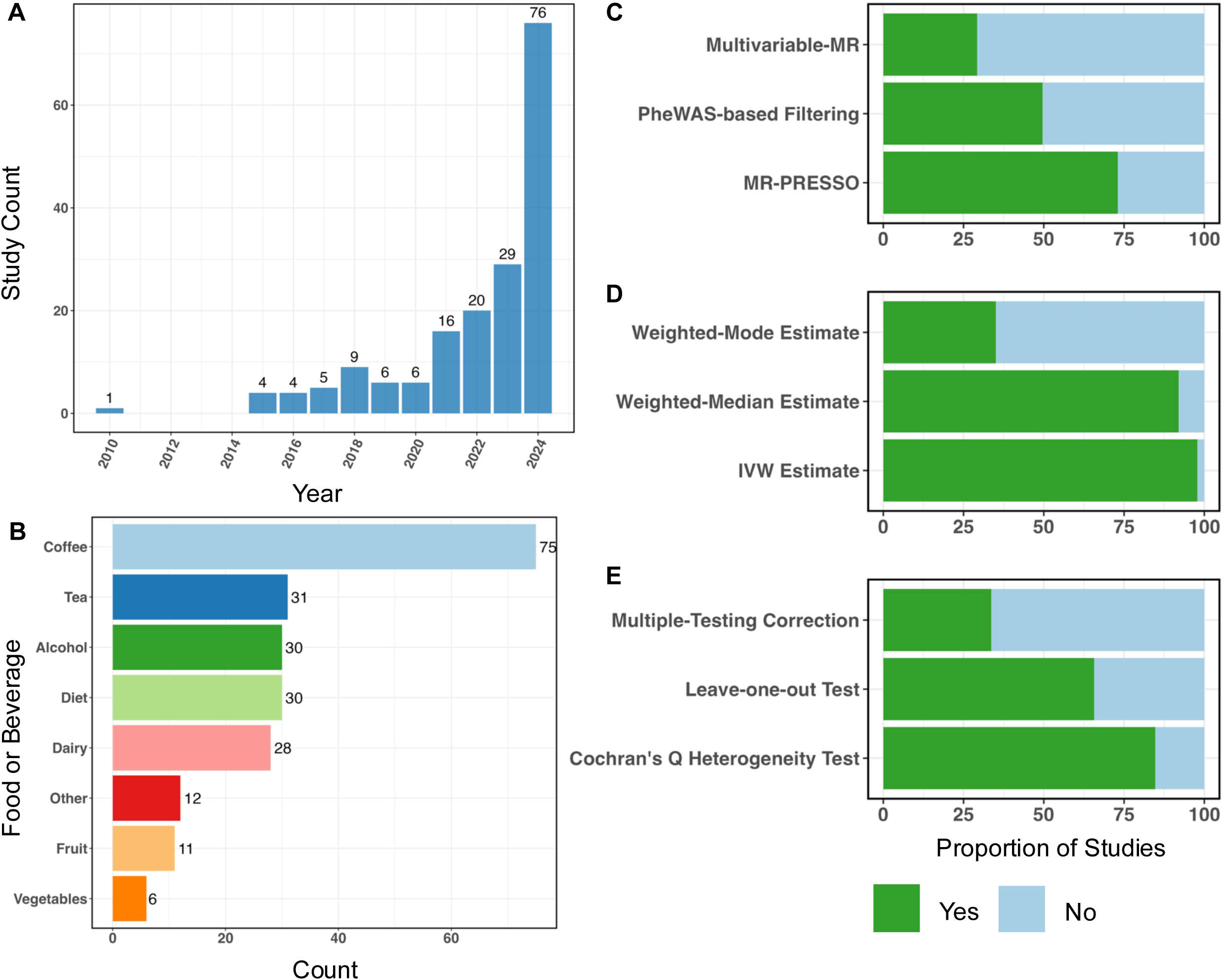
Assessment of the diet MR literature. **A**: Histogram showing diet MR studies published by year. When the search was conducted (Jan 24, 2025) one study was published in 2025 and is not pictured. **B**: Distribution of the foods and beverages studied. Studies evaluating more than 5 foods are included in “Diet.” “Other” included cereal, sugar-sweetened beverages, dark chocolate, noodles, meats, and nuts. **C**: The proportion of studies that used each method designed to address GI specificity. **D**: The proportion of studies that used each MR estimator. **E**: The proportion of studies that applied each sensitivity test.

The most common dietary exposure was coffee (n=75), followed by tea (n=31), alcohol (n=30), overall diet (n=30), and dairy (n=28) (**Figure 2B**). Most studies used GWS variants to build the GI (n=143, 80%), most often using the standard genome-wide significance threshold (5×10^−8^; n=110, 77%); however, a considerable number used a more permissive threshold, such as 5×10^−6^. GIs for thirty-five of the studies (20%) were composed of variants with a known functional relationship to the diet exposure. For example, dairy intake was commonly represented by rs4988235, which impacts the regulation of the lactase gene (*LCT*) and is associated with lactase persistence [33, 34]. Coffee and tea intake was often represented by variants in caffeine metabolizing or regulating genes such as cytochrome P450 family 1 subfamily a polypeptide 1/2 (*CYP1A1/2*) and aryl hydrocarbon receptor (*AHR*), but some studies also included GWS variants [35].

Most diet MR studies have been based on individuals with European ancestry (n=158, 89%), reflecting the biases in the availability of GWAS summary statistics. Additionally, no studies utilized sex-specific GIs for diet, even when using sex-specific genetic effects for outcomes such as ovarian or prostate cancer (n=22, 12%).

Most manuscripts considered instrument specificity in some fashion (**Figure 2C**). The most common method (n=105, 73%) was MR-PRESSO, which removes variants that are statistical outliers in the causal relationship. However, MR-PRESSO may result in false positives if the variants it removes to reduce heterogeneity are indeed driven by biology. In contrast, MVMR, which incorporates additional data to estimate the direct effect of diet while controlling for confounders, was only performed 28% of the time; the traits included in MVMR ranged from other dietary factors to biological traits relevant to the outcome. Socioeconomic and environmental factors like education and physical activity levels were rarely considered in MR, although they were often adjusted for in observational (non-genetic) analyses when studies performed both non-genetic and genetic analyses. Approximately half of the studies using a GWS GI used a PheWAS-based method (n=74, 52%), such as Phenoscanner [36] or searched the GWAS catalog, to check for pleiotropy. Studies employing PheWAS-based methods inconsistently described key methodological choices, including which traits were examined for pleiotropy, how those traits were chosen, and the thresholds used to exclude variants. Steiger filtering, which evaluates the suspected direction of the variant’s effect on the exposure vs. the outcome, was only performed in 6% of studies published after the method was established in 2017.

Among studies that used a GWS GI (n=143), the most common estimators included the inverse variance weighted (IVW; n=140, 98%) and weighted median (WM; n=135, 94%) estimators (**Figure 2D**). Most studies (n=123, 86%) evaluated instrument strength using the F-statistic, with the common threshold of F>10 considered reasonably strong. Sensitivity analyses, including the leave-one-out method (n=100, 70%) and calculating Cochran’s Q-statistic (n=127, 89%), were frequently used to evaluate heterogeneity (**Figure 2E**). Other methods to evaluate heterogeneity were cited less frequently, including the use of funnel plots (n=54, 38%).

### Diet-Health Associations

We then tested whether diet-MR using conventional two-sample MR approaches could detect relationships supported by nutrition science. The diet traits and their corresponding health outcomes and literature support are described in **Table 1**.

**Table 1.** Description of the positive and negative controls used in the MR analyses, including the nutrition science literature supporting the relationships.

| Dietary Exposure | Outcome | Expected Effect | Study Design and Reference |
| --- | --- | --- | --- |
| Oily Fish | TG | Decrease | RCT<sup>10</sup> |
|  | CVD | Decrease | RCT<sup>10</sup> |
|  | Height | None | NA |
| White vs Whole Grain or Brown Bread | LDL-C | Increase | RCT<sup>15</sup>, systematic review and meta-analysis<sup>16</sup> |
|  | CVD | Increase | Systematic review and meta-analysis<sup>17</sup> |
|  | Height | None | NA |
| Alcohol Intake | ALT | Increase | Prospective<sup>11</sup> |
|  | Liver Cirrhosis | Increase | Prospective<sup>13</sup>, animal<sup>14</sup> |
|  | Height | None | NA |

### Genome-wide significant genetic instruments do not consistently identify the expected relationships between diet-health positive controls

We performed MR using GIs consisting of GWS variants. The IVW estimate did not identify the expected relationship for any diet-health pairs (i.e., inconsistent direction or *P*>0.005) (**Figure 3A**). The relationship between alcohol and ALT was statistically significant, but in the opposite direction of what would be expected, such that each standard deviation (SD) increase in alcohol intake was associated with a decrease in ALT levels (ß=-0.02 [95% CI –0.035 to –0.008]). Steiger filtering was employed to select variants that acted in the desired causal direction (diet to health outcome) and tended to have a minimal effect on the GIs and MR estimates (**Table 2**). However, for white vs whole grain or brown bread on LDL-C, Steiger filtering removed one variant known to be involved in cholesterol metabolism (rs429358), rescuing the expected positive association (IVW: ß=0.1 [95% CI 0.05 to 0.15]). Although there was no evidence of weak instrument bias (F range: 36-41 for each positive control), the GIs showed heterogeneity (**Table 2**). The IVW method did not detect false positives amongst the negative control outcome of height (**Figure 3B**).

**Figure 3.**
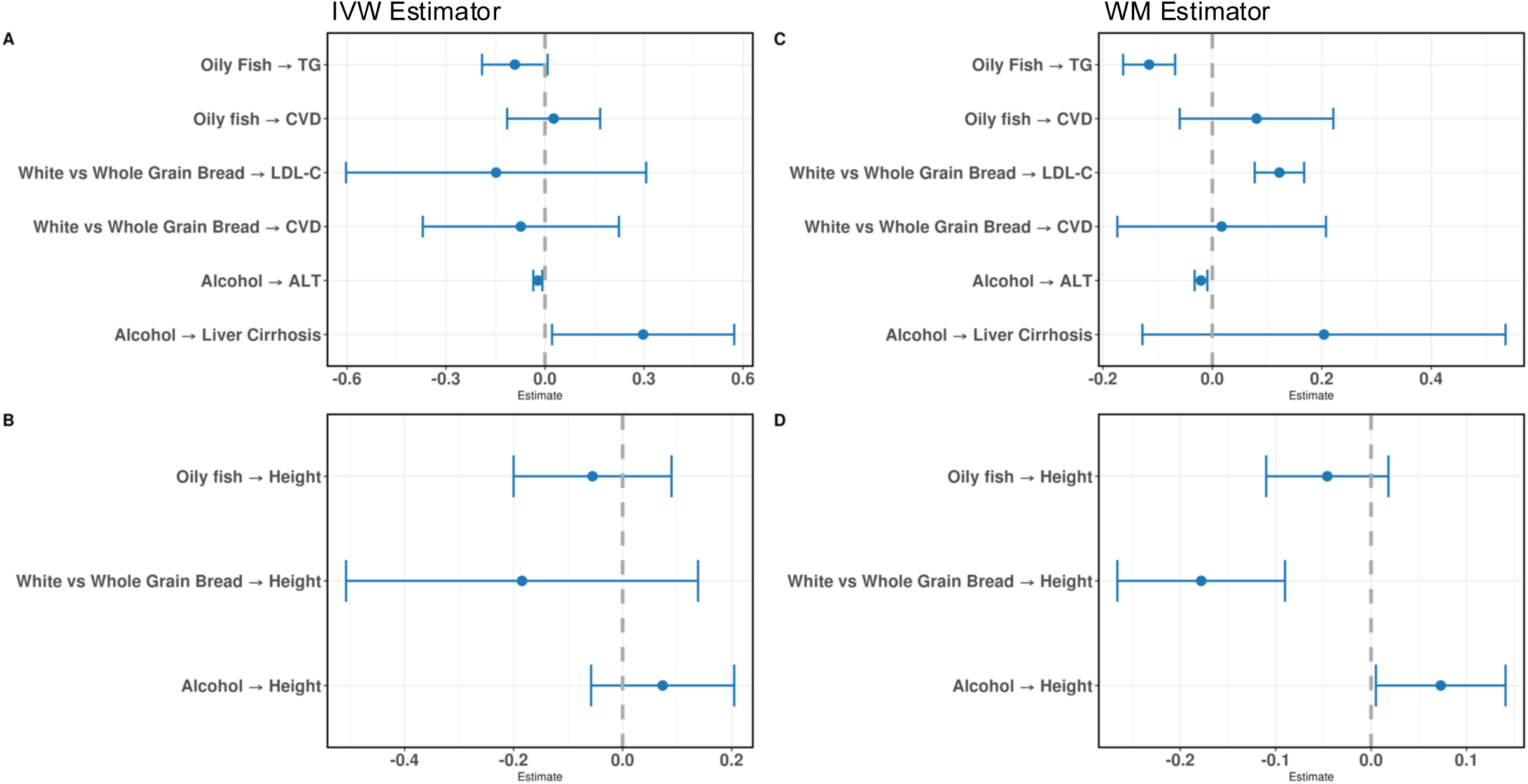
MR estimates from the GWS GI. **A:** The positive controls tested by the IVW estimator. **B:** The negative control tested by the IVW estimator. **C:** The positive controls tested by the WM estimator. **D:** The negative control tested by the WM estimator. The whiskers represent 95% confidence intervals. TG, triglycerides; LDL-C, LDL cholesterol; CVD, cardiovascular disease; ALT, alanine aminotransferase; IVW, inverse variance weighted; WM, weighted median.

**Table 2.**
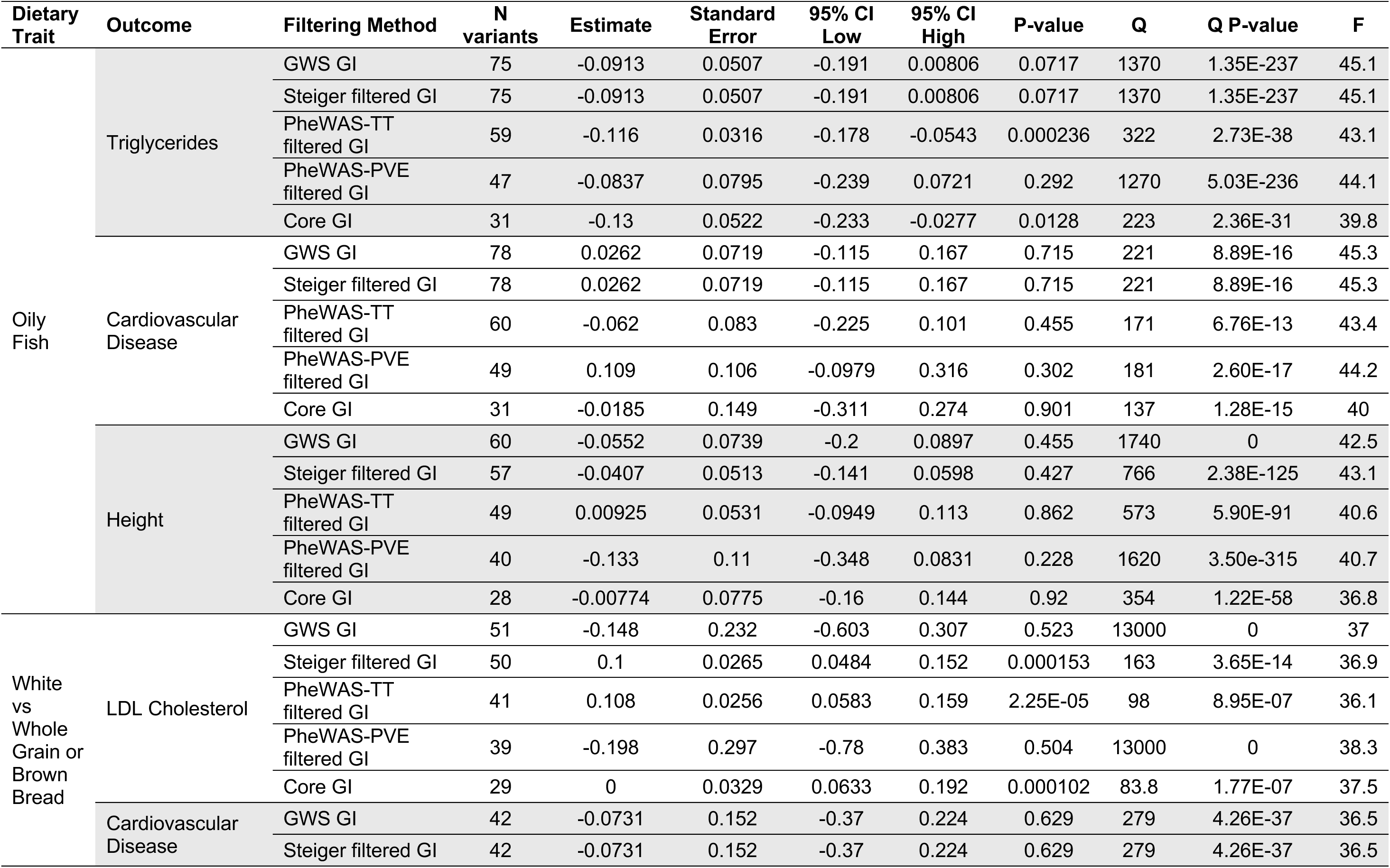
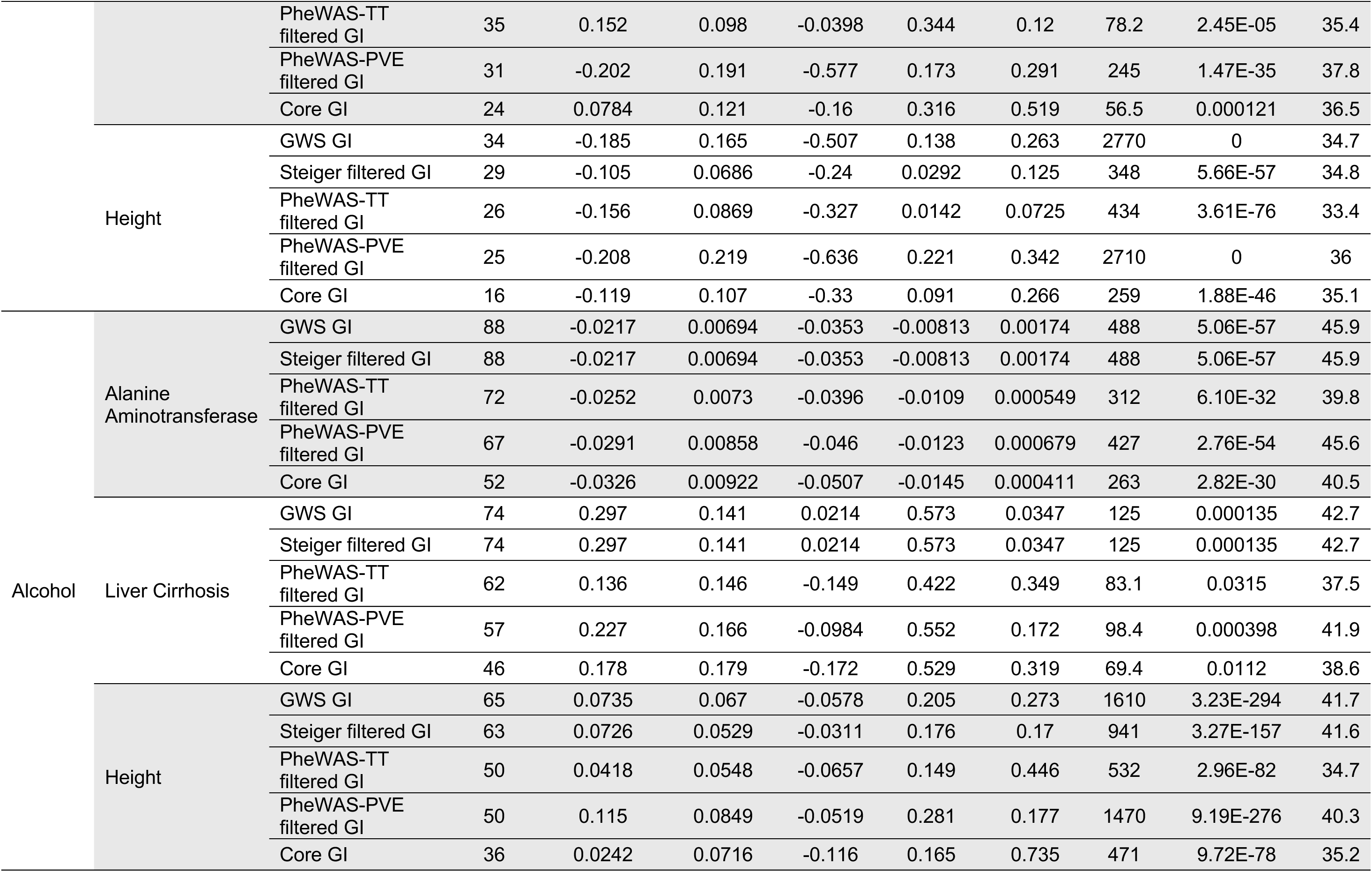
MR results calculated using the IVW estimator for the GWS and filtered GIs.

In contrast to the IVW method, the WM method, which is more robust to the exclusion restriction assumption by allowing at least 50% of the variants to be valid, identified statistically significant relationships between oily fish on triglycerides (ß=-0.12 [95% CI –0.16 to –0.07]) and bread type on LDL cholesterol levels (ß=0.12 [95% CI 0.08 to 0.17]) (**Figure 3C**). Like the IVW, the WM method estimated a statistically significant inverse effect of alcohol on ALT levels (ß =-0.02 [95% CI –0.03 to –0;01]). The WM method identified false positives, including white vs whole grain or brown bread on height (ß =-0.12, [95% CI –0.27 to –0.09]) and a nominal effect (P=0.03, not surpassing the multiple testing correction threshold) of alcohol on height (ß =0.073, [95% CI 0.005 to 0.14]) (**Figure 3D**). After Steiger filtering, the relationship between bread type on height was nullified (**Table 3**).

**Table 3.**
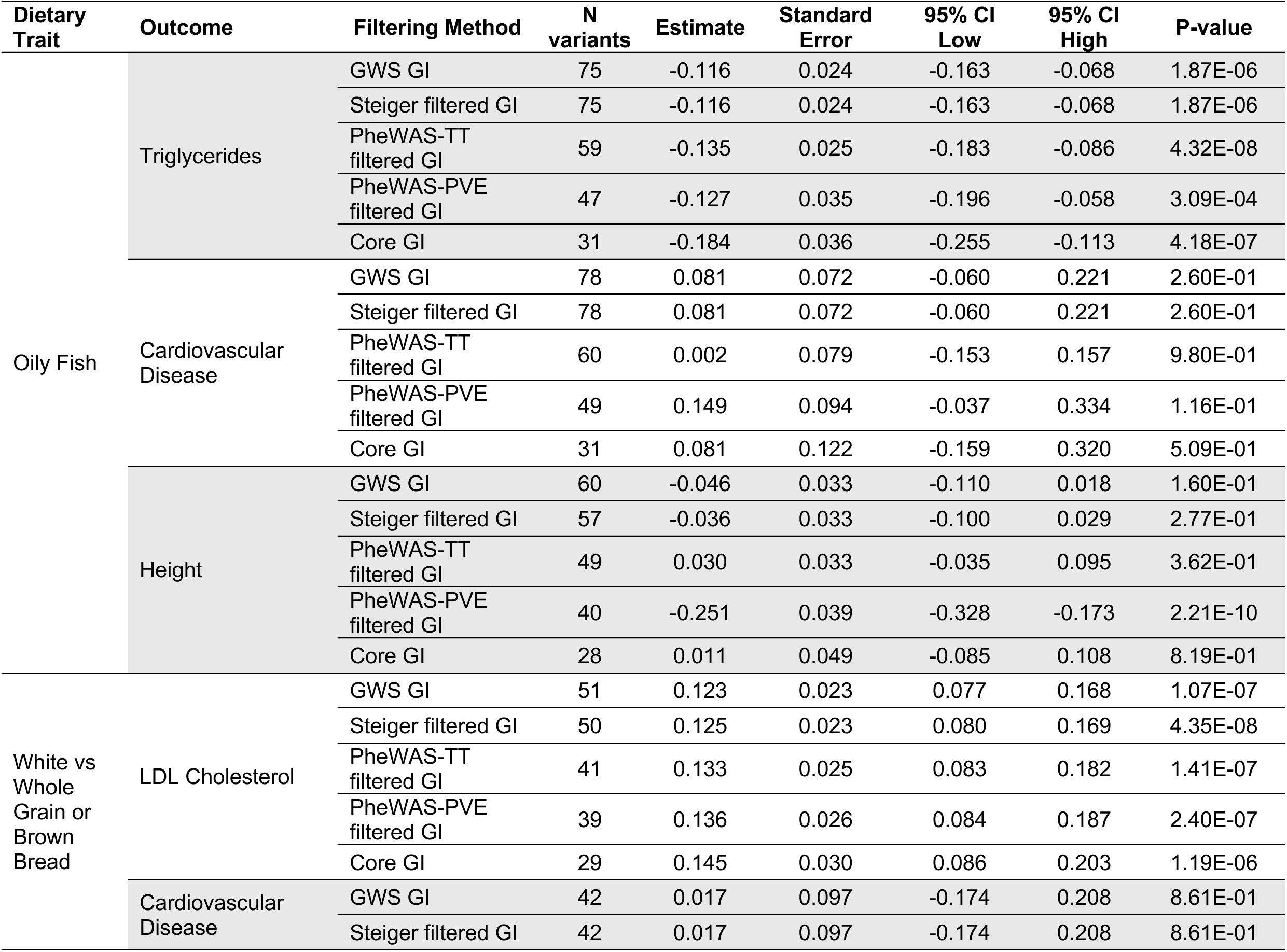
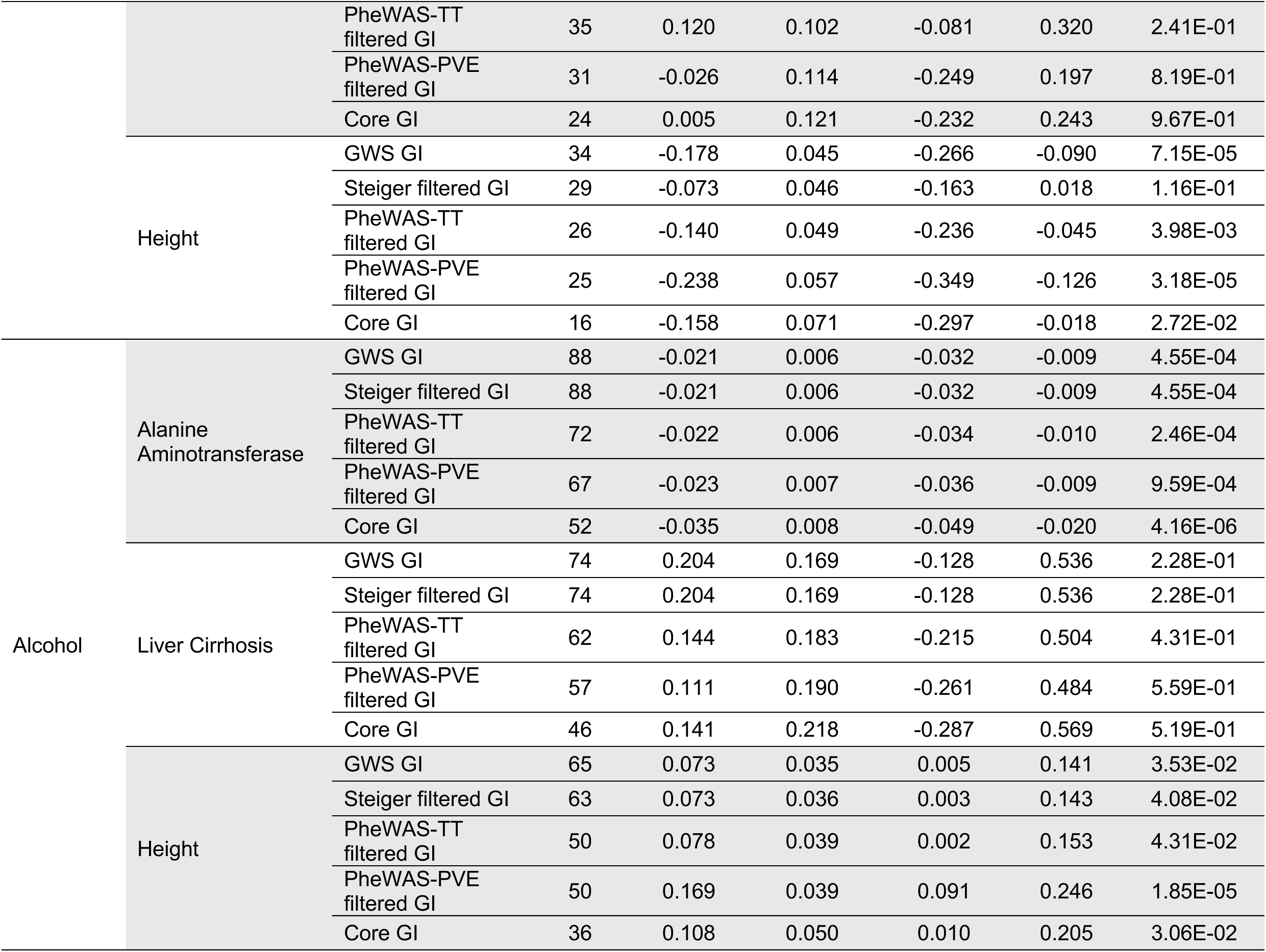
MR results calculated using the WM estimator for GWS and filtered GIs.

### MR-CAUSE

Due to the polygenic genetic architecture influencing dietary intake, we tested MR-CAUSE, a method that utilizes genome-wide effects as opposed to GWS effects. MR-CAUSE compares the fit of the data under two models: a causal (exposure → outcome) and a shared model driven by an unknown confounder (confounder → outcome) (**Figure S2**). MR-CAUSE supported a causal model for oily fish decreasing triglycerides (γ=0.08 [-0.11, –0.05]), white vs whole grain or brown bread increasing LDL-C (γ=0.05 [0.02, 0.07]), and CVD (γ=0.17 [0.09, 0.25]), and alcohol decreasing ALT (γ=-0.01 [-0.02, –0.01]) (**Table 4** and **5**). Comparing the null and shared models, MR-CAUSE also supported that these relationships were driven, in part, by an unknown confounder (**Table 4**). This contrasted with the findings of the commonly used MR-Egger intercept method which found no evidence of directional pleiotropy (P>0.075 in all cases, **Supplemental Table 4**). Although MR-CAUSE generally performed better than the GWS GIs, it falsely identified relationships between white vs. whole grain or brown bread (γ=-0.09 [-0.14, –0.04]) and alcohol on height (γ=0.07 [0.03, 0.11]) (**Table 5**).

**Table 4.** MR-CAUSE results demonstrating the fit of the shared and causal models for each positive and negative control. The statistics represent the comparison of model 1 and model 2.

| Dietary Trait | Outcome | Model 1 | Model 2 | ΔELPD | Standard Error of ΔELPD | Z | P-Value |
| --- | --- | --- | --- | --- | --- | --- | --- |
| Oily Fish | Triglycerides | Null | Sharing | -14.554 | 3.436 | -4.235 | 1.14E-05 |
|  |  | Null | Causal | -21.366 | 5.074 | -4.211 | 1.27E-05 |
|  |  | Sharing | Causal | -6.812 | 1.649 | -4.131 | 1.81E-05 |
|  | Cardiovascular Disease | Null | Sharing | 0.476 | 0.041 | 11.613 | 1 |
|  |  | Null | Causal | 1.263 | 0.083 | 15.198 | 1 |
|  |  | Sharing | Causal | 0.787 | 0.066 | 11.991 | 1 |
|  | Height | Null | Sharing | -0.060 | 0.461 | -0.130 | 0.448 |
|  |  | Null | Causal | -1.697 | 2.108 | -0.805 | 0.210 |
|  |  | Sharing | Causal | -1.636 | 1.656 | -0.988 | 0.161 |
| White vs Whole Grain or Brown Bread | LDL Cholesterol | Null | Sharing | -3.935 | 1.861 | -2.115 | 0.017 |
|  |  | Null | Causal | -8.399 | 3.818 | -2.200 | 0.014 |
|  |  | Sharing | Causal | -4.464 | 1.970 | -2.266 | 0.012 |
|  | Cardiovascular Disease | Null | Sharing | -5.505 | 2.356 | -2.337 | 0.010 |
|  |  | Null | Causal | -10.128 | 4.152 | -2.439 | 0.007 |
|  |  | Sharing | Causal | -4.624 | 1.830 | -2.527 | 0.006 |
|  | Height | Null | Sharing | -2.060 | 1.253 | -1.644 | 0.050 |
|  |  | Null | Causal | -6.385 | 3.246 | -1.967 | 0.025 |
|  |  | Sharing | Causal | -4.325 | 2.005 | -2.157 | 0.016 |
| Alcohol | Alanine Aminotransferase | Null | Sharing | -8.006 | 2.964 | -2.701 | 0.003 |
|  |  | Null | Causal | -12.544 | 4.708 | -2.664 | 0.004 |
|  |  | Sharing | Causal | -4.538 | 1.774 | -2.558 | 0.005 |
|  | Liver Cirrhosis | Null | Sharing | 0.424 | 0.302 | 1.403 | 0.920 |
|  |  | Null | Causal | 1.061 | 0.730 | 1.453 | 0.927 |
|  |  | Sharing | Causal | 0.637 | 0.440 | 1.447 | 0.926 |
|  | Height | Null | Sharing | -2.312 | 1.524 | -1.517 | 0.065 |
|  |  | Null | Causal | -5.475 | 3.327 | -1.646 | 0.050 |
|  |  | Sharing | Causal | -3.163 | 1.834 | -1.724 | 0.042 |

**Table 5.** MR-CAUSE results demonstrating the estimated causal effect (γ), shared effect (η), and uncorrelated effect (q) in the shared and causal models for each positive and negative control. In the shared model, the causal effect is set to 0.

| Dietary Trait | Outcome | Model | Gamma, $\gamma$ | Eta, $\eta$ | $q$ |
| --- | --- | --- | --- | --- | --- |
| Oily fish | Triglycerides | Sharing | NA | -0.11 (-0.17, -0.07) | 0.49 (0.24, 0.71) |
|  |  | Causal | -0.08 (-0.11, -0.05) | 0 (-0.31, 0.3) | 0.03 (0, 0.25) |
|  | Cardiovascular Disease | Sharing | NA | -0.01 (-0.79, 0.79) | 0.03 (0, 0.22) |
|  |  | Causal | 0 (-0.07, 0.06) | 0 (-0.72, 0.72) | 0.04 (0, 0.25) |
|  | Height | Sharing | NA | 0.14 (-0.33, 0.79) | 0.05 (0, 0.31) |
|  |  | Causal | 0.05 (0, 0.09) | 0.01 (-0.57, 0.69) | 0.03 (0, 0.25) |
| White vs Whole Grain or Brown Bread | LDL Cholesterol | Sharing | NA | 0.1 (0.03, 0.22) | 0.23 (0.02, 0.51) |
|  |  | Causal | 0.05 (0.02, 0.07) | 0 (-0.36, 0.31) | 0.03 (0, 0.25) |
|  | Cardiovascular Disease | Sharing | NA | 0.37 (0.17, 0.75) | 0.27 (0.05, 0.53) |
|  |  | Causal | 0.17 (0.09, 0.25) | -0.01 (-0.91, 0.84) | 0.04 (0, 0.25) |
|  | Height | Sharing | NA | -0.2 (-0.61, 0.02) | 0.17 (0.01, 0.46) |
|  |  | Causal | -0.09 (-0.14, -0.04) | 0 (-0.63, 0.6) | 0.03 (0, 0.25) |
| Alcohol | Alanine Aminotransferase | Sharing | NA | -0.02 (-0.04, -0.01) | 0.31 (0.09, 0.55) |
|  |  | Causal | -0.01 (-0.02, -0.01) | 0 (-0.14, 0.09) | 0.03 (0, 0.24) |
|  | Liver Cirrhosis | Sharing | NA | 0.32 (-1.2, 2.48) | 0.03 (0, 0.22) |
|  |  | Causal | 0.03 (-0.1, 0.15) | 0.11 (-1.29, 1.86) | 0.04 (0, 0.25) |
|  | Height | Sharing | NA | 0.19 (0.01, 0.56) | 0.16 (0.01, 0.43) |
|  |  | Causal | 0.07 (0.03, 0.11) | 0.01 (-0.59, 0.61) | 0.03 (0, 0.25) |

### Instruments are imprecisely specified, and systematic filtering of the genetic instrument can improve the detection of diet and health outcome positive controls

Due to the inconsistent findings of the GWS GIs, we then tested if systematically filtering the GI using PheWAS or controlling for confounding via MVMR would better identify the expected relationships. A comparison of the statistical assumptions of the conventional and robust MR estimators is given in **Table 6**.

**Table 6.**
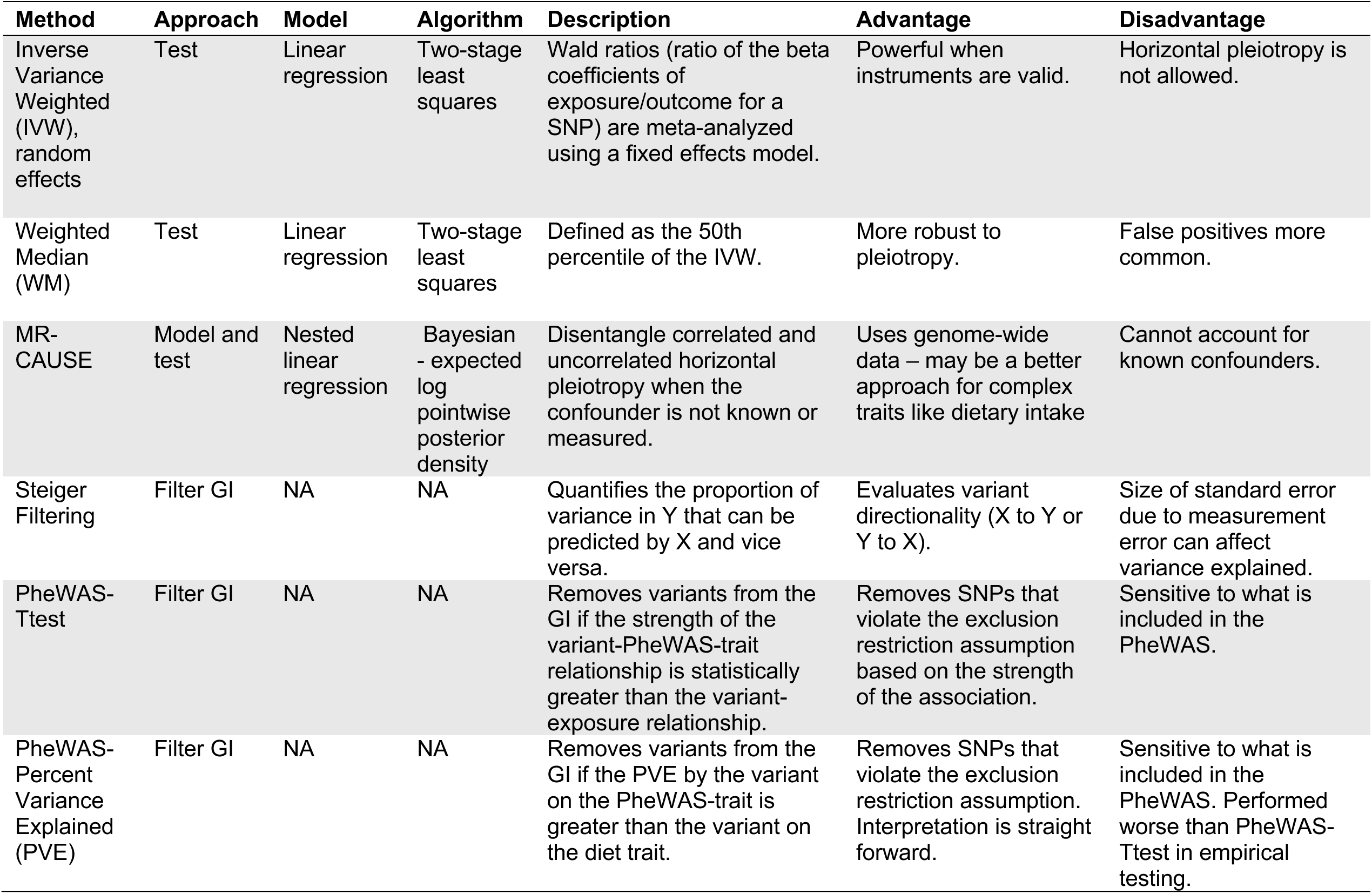
Descriptions of the MR methods used in the positive and negative control analysis.

MR estimates derived from GIs filtered by the PheWAS-Ttest method performed better than the PheWAS-PVE method (**Figure 4**). The PheWAS-Ttest GI filtering method identified the expected causal effects between oily fish on TG (IVW: ß=-0.12 [95% CI –0.18 to –0.054]; WM: ß=-0.14 [95% CI –0.18 to –0.086]) and white vs. whole grain or brown bread on LDL-C (IVW: ß = 0.11 [95% CI 0.058 to 0.16]; WM: ß = 0.13 [95% CI 0.083 to 0.18]) via the IVW and WM estimators, whereas the PheWAS-PVE filtering method only identified significant relationships when using the WM estimator (oily fish on TG, WM: ß=-0.13 [95% CI –0.20 to –0.058]; white vs whole grain or brown bread on LDL-C, WM: ß=0.14 [95% CI 0.084 to 0.19]). The PheWAS-Ttest method tended to retain more variants while limiting GI heterogeneity versus the PheWAS-PVE method (**Table 2**). Variants and their associated traits removed by the PheWAS-Ttest and PheWAS-PVE methods were dominated by anthropometric measurements but included some social determinants of health such as level of education (**Supplemental Tables 5** and **6**, respectively).

**Figure 4.**
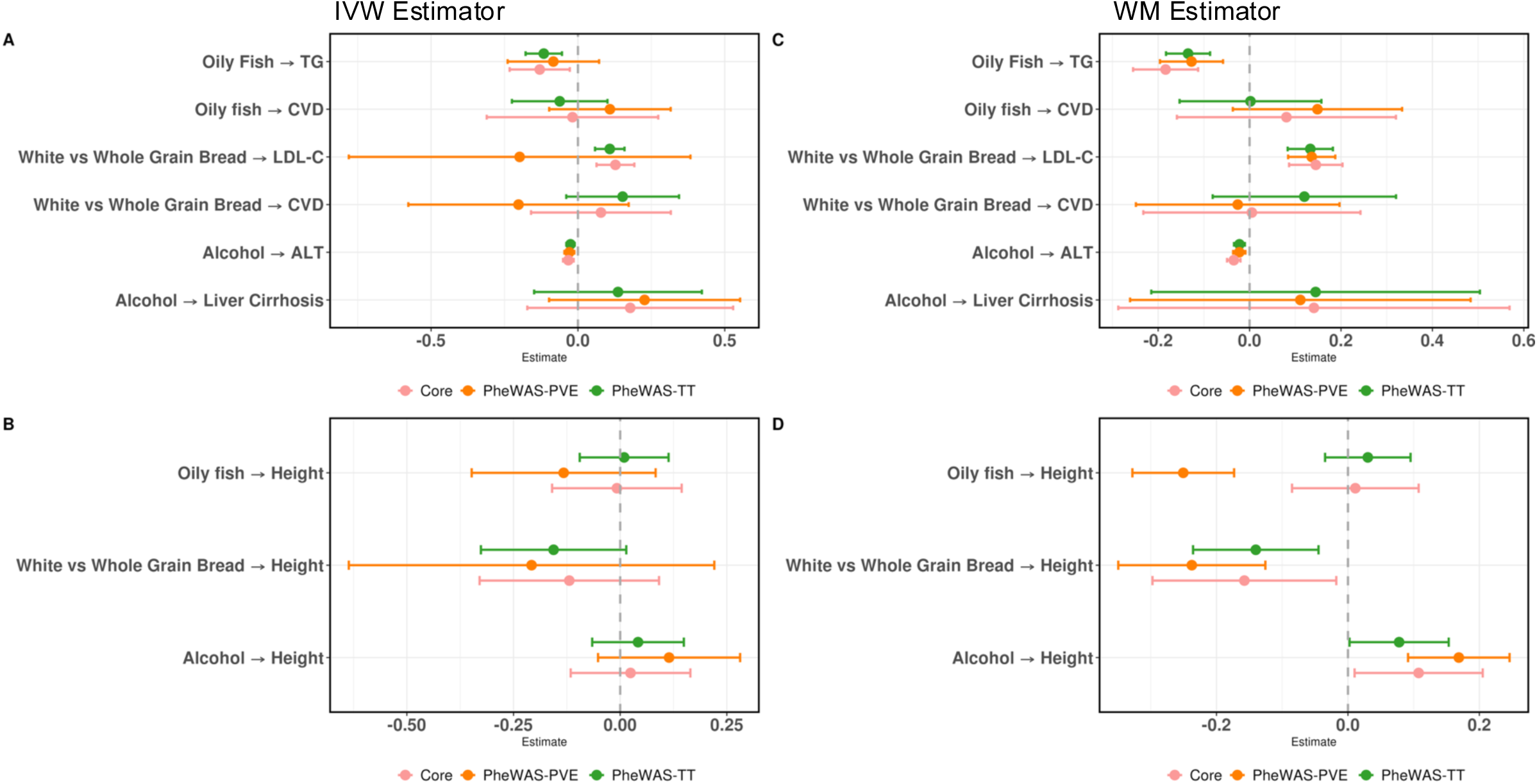
MR estimates from the GI filtered using the PheWAS-Ttest (green), PheWAS-PVE (orange), or Core (pink) methods. **A:** The positive controls tested by the IVW estimator. **B:** The negative control control by the IVW estimator. **C:** The positive controls tested by the WM estimator. **D:** The negative tested by the WM estimator. The whiskers represent 95% confidence intervals. TG, triglycerides; LDL-C, LDL cholesterol; CVD, cardiovascular disease; ALT, alanine aminotransferase; IVW, inverse variance weighted; WM, weighted median.

Due to the substitutional nature of dietary intake, we performed a sensitivity PheWAS analysis excluding any dietary traits. Nevertheless, the MR results for the IVW (**Supplemental Table 7**) and WM tests (**Supplemental Table 8**) remained stable for both the PheWAS-Ttest and PheWAS-PVE methods.

We calculated the causal estimates using only the variants that were retained following Steiger filtering and both PheWAS approaches (coined “core” GI). For the positive controls, the core GI performed similarly to the PheWAS-Ttest method. No false positives were detected by the core GIs (**Tables 2** and **3**). The effects of all filtering approaches, including the core approach, are presented in **Figure 4**. Regardless of the GI filtering method, no significant relationships were detected between diet traits and diseases.

### PheWAS informed trait selection for MVMR

Because the PheWAS-Ttest method performed closer to the literature expectation than the PheWAS-PVE method, we developed a pipeline to objectively identify traits for MVMR using the PheWAS-Ttest output. The MVMR pipeline demonstrated the importance of controlling for confounding traits and rescued the expected positive causal estimate between alcohol and ALT. Prior to MVMR, the GWS GI and filtered GIs consistently identified a significant inverse relationship between alcohol intake on ALT. The MVMR pipeline identified five clusters that confounded the alcohol GIs. Anthropometric traits dominated the clusters, which were represented by platelet count, blood volume, weight, sitting height, and body impedance. Accounting for these traits resulted in a direct effect of alcohol on ALT (IVW: ß= 0.028 [95% CI 0.017 to 0.039]) as expected from the literature (**Table 7**). The direct effects of weight (IVW: ß= –0.027 [95% CI –0.035 to –0.019]) and sitting height (IVW: ß= 0.019 [95% CI 0.013 to 0.024]) were also significant. Although the expected relationship was identified, the GI was large, containing over 2000 independent variants (dominated by the anthropometric traits), with significant heterogeneity (Q=10116, P<0.005) and varying F-statistics (range: 6-98). We then analyzed the remaining diet-health relationships with MVMR (**Supplemental Table 9**) and found the oily fish on TG association remained marginally significant after adjusting for predominantly blood-related confounding traits (IVW: ß= 0.14 [95% CI 0.007 to 0.27]).

**Table 7.**
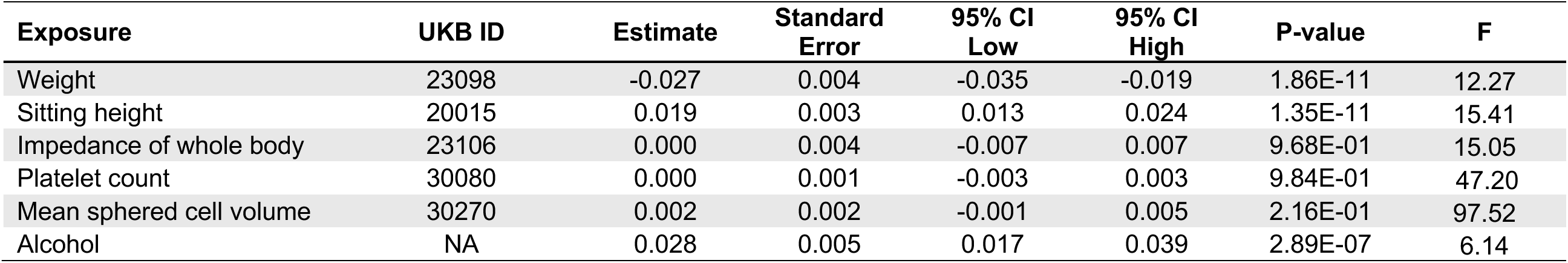
MVMR results for alcohol on ALT levels using additional exposures identified from the PheWAS-Ttest pipeline.

## Discussion

First, we performed an assessment of the diet-MR literature to outline the overarching applications, common practices, and considerations in the field. Diet-MR analyses are flooding the literature with a wide range of methodological rigor with respect to the core MR assumptions. We then tested whether conventional MR tests using GWS GI could identify relationships between dietary exposures and their established health outcomes using only publicly available GWAS data. Due to the myriad factors known to influence dietary intake and previous evidence showing extensive genetic correlations between diet and other behavioral factors [5, 37], we hypothesized that many of the variants within the GWS GI affected dietary intake through indirect, pleiotropic paths, violating the exclusion restriction assumption. Thus, we evaluated whether systematically filtering the GI would improve the ability to detect the literature-supported positive controls by capturing the direct effect of diet on health. Finally, we used MVMR, coupled with a pipeline to systematically identify mediators for inclusion, to estimate the direct causal effects of diet on health.

### Findings from the literature assessment

The availability of GWAS summary statistics and the software to perform MR has contributed to the rapid rise of MR studies, leading to concern about the credibility of the approach [38]. The Strengthening the Reporting of Observational Studies in Epidemiology (STROBE) guidelines were used to develop the STROBE-MR guidelines, which serve as a checklist to promote thorough analyses and reporting of MR studies [39]; however, their uptake has been modest. Other guidelines for performing MR have been published, but have not been sufficient to prevent poorly conducted studies [40]. Our main concerns are described below.

*GI specificity.* Socioeconomic and behavioral factors like income, education level, and physical activity levels, as well as body composition and anthropometrics, are genetically and phenotypically correlated with dietary intake with varying overlap at GWS loci. However, many MR studies did not consider how these relationships influenced the causal estimates. Furthermore, most diet-MR studies to date have been conducted using GIs from GWAS of European ancestry of dietary intake traits derived from a 29-question FFQ or a single >200-nested-question 24-hour recall from the UKB. Although a valuable resource, oftentimes these GWAS do not consider information from multiple diet recalls or extreme intakes, and therefore, researchers performing diet-MR should consider whether the underlying diet GWAS phenotype captured the diet trait reported in their MR.

*Appropriate application of MR.* Applying MR indiscriminately on numerous diet traits and diseases in a hypothesis-free manner is a questionable practice. GIs whose biological function impacts the intake of a given food, such as variants that impact lactase persistence or caffeine metabolism, are considered the gold standard, though unfortunately often have lower power than GWS GI. However, the function of most GWS variants is unknown, making it especially important to assess how the GWAS data were derived, whether the instruments are associated with confounders, and then vet the causal estimates with numerous estimators and sensitivity tests.

*Common methods observed in diet-MR.* MR-PRESSO was the most common method for filtering variants from the GI that were suspected to violate the exclusion-restriction assumption. This outlier-removal method has a high false positive rate if many of the instruments are invalid, which is likely prevalent in diet MR. Phenoscanner, an online tool facilitating phenome scans, was the most cited tool for filtering variants based on instrument specificity, but it is no longer publicly accessible. In its absence, applying this principle using the GWAS catalog or PheWAS-based approaches on UKB or related datasets can help address GI specificity. Many studies combined multiple MR estimators, including the IVW, weighted median, and MR-Egger tests, which should continue in addition to using newer robust methods, and those that particularly allow for causal and shared paths (MR-CAUSE) or direct and indirect effects (MVMR).

### Findings from the positive and negative control analyses

Despite displaying acceptable values of instrument strength, the GWS GIs displayed significant heterogeneity and largely failed to identify the expected diet-health relationships. Filtering the GI using Steiger filtering, the PheWAS methods, or the core method better captured the expected diet-outcome relationships, but each method had false positives and/or negatives. Notably, the Steiger filtering removed variants that had a strong effect on the outcome, whereas the PheWAS methods tended to remove variants that had strong associations with confounding traits, which are not tested by existing outlier-based methods. These results reinforce the existing recommendation to base interpretations on the combined evidence of several methods [39].

Despite epidemiological evidence linking dietary factors to diseases, our MR analyses only identified relationships with intermediate biomarkers of disease and not the diseases themselves. Biomarkers precede health diagnosis and are often one indicator of many in receiving a disease diagnosis. The failure of our MR to identify diet-disease relationships may be due to the complexity of the factors that influence disease status and perhaps focusing on intermediate biomarkers and underlying nutrients for dietary exposures will resolve more causal relationships.

### Sources of confounding

Individuals who receive medical information about their health may alter their diet in response. The variant rs429358 was positively associated with both consuming whole grain or brown bread and LDL cholesterol. The variant is in the fourth exon of the *ApoE* gene and the effect allele (C) has an established association with LDL-C levels [41].

Individuals with the risk allele may have high LDL-C due to their genetics and therefore (unknowing of their genotype) be more likely to consume fiber-rich bread sources to reduce their cholesterol. Whether diet drives health behaviors or vice versa can be explored using bi-directional MR. A similar phenomenon may explain the counterintuitive relationship between alcohol and ALT. Although ALT levels are affected by alcohol intake, many have reported that this is mediated by body mass index (BMI), such that the effect is observed at high but not normal BMI [12]. We investigated this using MVMR and identified that alcohol indeed increased ALT levels when controlling for anthropometric factors.

Others have raised the concern that genetic loci associated with dietary intake are partly driven by indirect effects from confounding traits and developed nuanced workflows requiring individual-level data to address this [37]. For example, MR GIs have been identified using a multi-step approach described by first performing GWAS using a Bayesian estimation to account for the effect of other traits on dietary intake. And second, including variants in the GI that are statistically supported to have a direct effect on diet based on the corrected to uncorrected ratio (CUR), a measurement of the magnitude of change of the effect when covariates are included in the analyses [42]. This effort revealed that the effect sizes of loci associated with diet are altered when adjusting for the confounders, such as BMI and education level, implying that these factors should be considered when estimating the effects of diet on health with MR.

Although we hypothesized that social determinants of health (SDOH) would confound the diet GIs, the pipeline mostly identified anthropometric-related traits. This finding may be due to the unequal representation of SDOH traits in the PheWAS data or the precision with which anthropometric traits are measured versus environmental factors like diet. Nevertheless, it is important to point out that traits that are genetically correlated such as diet and SDOH may not be strongly associated with the top diet loci.

### Limitations

There are several limitations of this work. Due to linkage disequilibrium, minor allele frequency, and data availability, these analyses were restricted to GWAS conducted in individuals with European genetic ancestry. Additionally, culturally appropriate dietary questionnaires in populations with different ancestry backgrounds would likely result in the use of a different FFQ, making the harmonization of phenotypes for future meta-analyses difficult. We originally sought to compare the effects of the lifelong genetically predicted exposure in MR to RCT studies in shorter windows. However, the ordinal nature of the diet traits, coupled with the difficulty of identifying RCTs solely testing the food of interest, made this goal unachievable.

### Future Directions for MR of Diet

Despite facing challenges, MR is a promising method for vetting observational findings to prioritize limited resources towards RCTs with a greater likelihood of success. To accomplish this, we have several suggestions for the future directions of diet-MR, including sex-aware and ancestry-diverse analyses, improving diet assessments in biobanks, and acknowledging the limitations of the original input data.

Both dietary intake and metabolic diseases have sex-specific differences, yet most GWAS have been conducted controlling for sex, likely missing numerous loci influencing these traits. Advances in genome-wide association testing have made genome-wide interaction studies by sex feasible, which will unlock new sex-aware MR analyses that will better capture the effect of diet on health by sex. Testing which foods and beverages are causally linked to health outcomes by sex and in other ancestries and cultures is important for personalization and generalizability.

Efforts to improve the accuracy and detail of dietary assessments are underway. Machine learning algorithms powered by databases of annotated food pictures are a promising technology that could be delivered at biobank-level scale to improve dietary phenotyping, resulting in a more precise understanding of the genetic architecture of diet [43–45]. Furthermore, the identification of molecular food biomarkers could serve to validate existing dietary assessments or could be leveraged as GIs that represent true dietary intake better than existing recall methods.

As the field progresses towards these methodological advancements, immediate attention should be given to the specificity of the GI, the source of the GWAS diet data, and the relevance of MR to address the diet-health question. In addition to recommendations from several general MR best-practices reviews [39, 40], researchers interested in diet-MR should vet the GI for pleiotropy/confounding using PheWAS-based methods such as the PheWAS-Ttest method and control for these factors using MVMR. Whenever possible, the GI should employ functional variants with biological relevance to the dietary trait.

### Conclusion

Concern about the credibility of diet-MR is timely due to the growing interest in this field. Due to the inconsistency and inaccuracy of the positive and negative controls across multiple MR estimators, MR using GIs derived from food frequency questionnaires or 24-hour dietary assessments should be used and interpreted with caution. Scrutiny of the GI is paramount to avoid mischaracterizing the causal relationships between dietary intake and health and further eroding public trust in nutrition science.

## Supporting information

STROBE-MR Checklist

## Data Availability

Data described in the manuscript are publicly and freely available without restriction at the GWAS catalog (Supplemental Table 1: LDL-C, GCST90239658; CVD, GCST90132314; TG, GCST90239664; ALT, GCST90013405; Cirrhosis of liver, GCST90013405; Height, GCST006901) and the Type 2 Diabetes Knowledge Portal by the dataset name, UK Biobank dietary habit GWAS.

## Abbreviations

ALT: alanine aminotransferase
CVD: cardiovascular disease
GI: genetic instrument
GWAS: genome-wide association study
IVW: inverse variance weighted
LDL-C: low density lipoprotein cholesterol
MeSH: medical subject headings
MR: Mendelian randomization
PheWAS: phenome-wide association study
PVE: percent variance explained
STROBE: strengthening the reporting of observational studies in epidemiology
TG: triglyceride
Ttest: t-test
WM: weighted median

## Acknowledgements

KJS, JEG, KW, DG, DH, AH and JBC designed research; KJS conducted research; KJS and MJ analyzed data; KJS wrote the first draft of the paper; KJS, JEG, KW, JBC edited the paper. KJS had primary responsibility for final content. All authors read and approved the final manuscript.

---

**Supplemental Figure 1.**
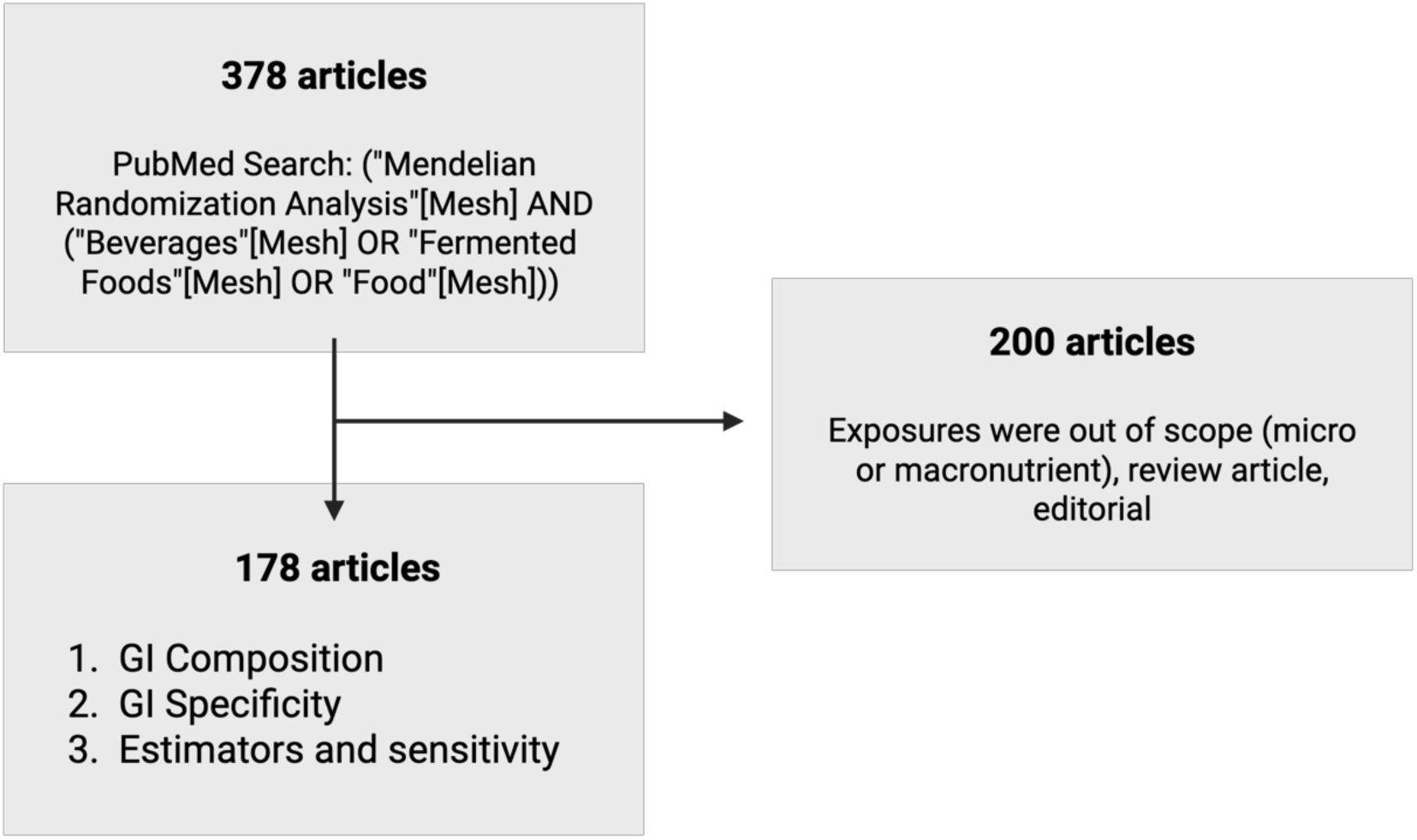
Diagram of the studies identified and included from the assessment of the dietary intake MR literature up to January 24, 2025. Mesh, Medical Subject Headings; GI, genetic instrument.

**Supplemental Figure 2.**
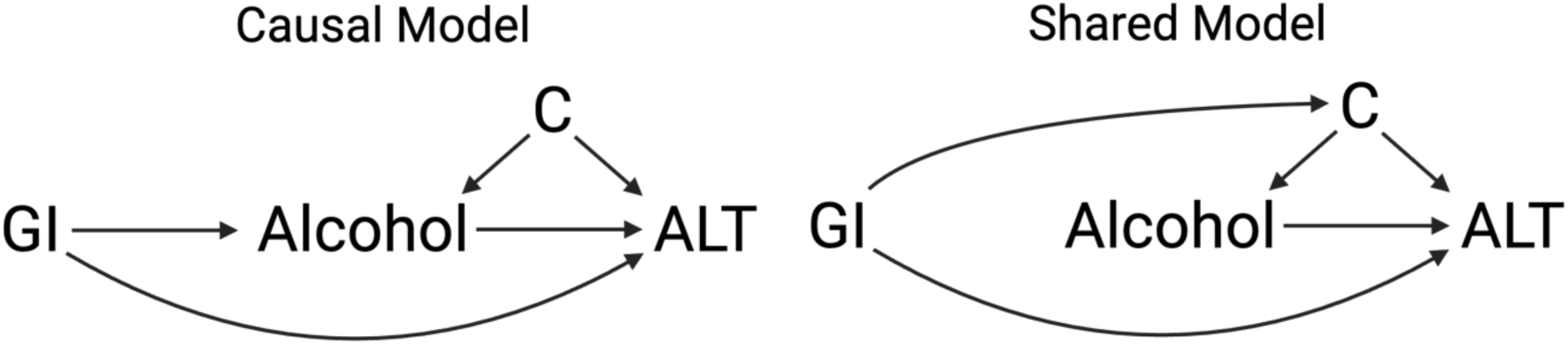
Example models demonstrating the causal and shared models from MR-CAUSE. C, confounder.

**Supplemental Figure 3.**
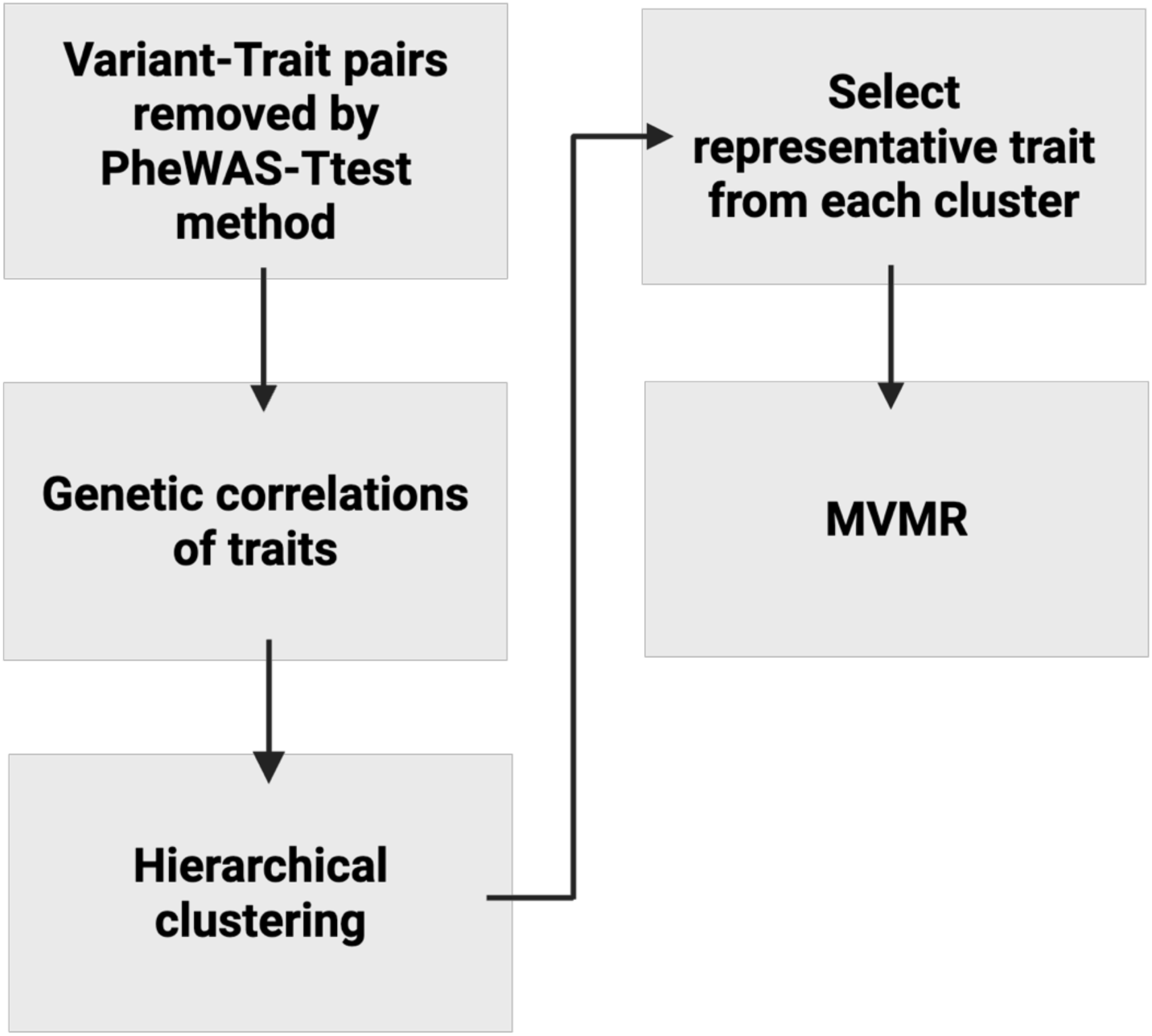
Illustration of the pipeline that uses the output of the PheWAS-Ttest method to identify traits to include in MVMR mediation analysis to tease apart diet’s direct and indirect paths on relevant health outcomes. The pipeline uses summary-level data, genetic correlation analysis, and hierarchical clustering to systematically select representative traits. MVMR, multivariable MR.

## References

[1] Klein EA, Thompson IM, Tangen CM, Crowley JJ, Lucia MS, Goodman PJ, Minasian LM, Ford LG, Parnes HL, Gaziano JM, et al. Vitamin E and the Risk of Prostate Cancer: The Selenium and Vitamin E Cancer Prevention Trial (SELECT). JAMA. 2011;306(14):1549. doi:10.1001/jama.2011.1437

[2] Homocysteine Lowering with Folic Acid and B Vitamins in Vascular Disease. New England Journal of Medicine. 2006;354(15):1567–1577. doi:10.1056/NEJMoa060900

[3] Sanderson E, Glymour MM, Holmes MV, Kang H, Morrison J, Munafò MR, Palmer T, Schooling CM, Wallace C, Zhao Q, et al. Mendelian randomization. Nature Reviews Methods Primers. 2022;2(1):6. doi:10.1038/s43586-021-00092-5

[4] May-Wilson S, Matoba N, Wade KH, Hottenga J-J, Concas MP, Mangino M, Grzeszkowiak EJ, Menni C, Gasparini P, Timpson NJ, et al. Large-scale GWAS of food liking reveals genetic determinants and genetic correlations with distinct neurophysiological traits. Nature Communications. 2022;13(1):2743. doi:10.1038/s41467-022-30187-w

[5] Cole JB, Florez JC, Hirschhorn JN. Comprehensive genomic analysis of dietary habits in UK Biobank identifies hundreds of genetic associations. Nature Communications. 2020;11(1):1467. doi:10.1038/s41467-020-15193-0

[6] Millard LA, Davies NM, Gaunt TR, Davey Smith G, Tilling K. Software Application Profile: PHESANT: a tool for performing automated phenome scans in UK Biobank. International Journal of Epidemiology. 2018;47(1):29–35. doi:10.1093/ije/dyx204

[7] Meddens SFW, De Vlaming R, Bowers P, Burik CAP, Linnér RK, Lee C, Okbay A, Turley P, Rietveld CA, Fontana MA, et al. Genomic analysis of diet composition finds novel loci and associations with health and lifestyle. Molecular Psychiatry. 2021;26(6):2056–2069. doi:10.1038/s41380-020-0697-5

[8] Verbanck M, Chen C-Y, Neale B, Do R. Detection of widespread horizontal pleiotropy in causal relationships inferred from Mendelian randomization between complex traits and diseases. Nature Genetics. 2018;50(5):693–698. doi:10.1038/s41588-018-0099-7

[9] Burgess S, Thompson SG. Multivariable Mendelian Randomization: The Use of Pleiotropic Genetic Variants to Estimate Causal Elects. American Journal of Epidemiology. 2015;181(4):251–260. doi:10.1093/aje/kwu283

[10] Bhatt DL, Miller M, Brinton EA, Jacobson TA, Steg PhG, Ketchum SB, Doyle RT, Juliano RA, Jiao L, Granowitz C, et al. REDUCE-IT USA: Results From the 3146 Patients Randomized in the United States. Circulation. 2020;141(5):367–375. doi:10.1161/CIRCULATIONAHA.119.044440

[11] Alatalo PI, Koivisto HM, Hietala JP, Puukka KS, Bloigu R, Niemelä OJ. Elect of moderate alcohol consumption on liver enzymes increases with increasing body mass index. The American Journal of Clinical Nutrition. 2008;88(4):1097–1103. doi:10.1093/ajcn/88.4.1097

[12] Ruhl CE, Everhart JE. Joint Elects of Body Weight and Alcohol on Elevated Serum Alanine Aminotransferase in the United States Population. Clinical Gastroenterology and Hepatology. 2005;3(12):1260–1268. doi:10.1016/S1542-3565(05)00743-3

[13] Simpson RF, Hermon C, Liu B, Green J, Reeves GK, Beral V, Floud S. Alcohol drinking patterns and liver cirrhosis risk: analysis of the prospective UK Million Women Study. The Lancet Public Health. 2019;4(1):e41–e48. doi:10.1016/S2468-2667(18)30230-5

[14] Lieber CS, Jones DP, DeCarli LM. Elects of Prolonged Ethanol Intake: Production of Fatty Liver Despite Adequate Diets*. Journal of Clinical Investigation. 1965;44(6):1009–1021. doi:10.1172/JCI105200

[15] Madsen MTB, Landberg R, Nielsen DS, Zhang Y, Anneberg OMR, Lauritzen L, Damsgaard CT. Elects of wholegrain compared to refined grain Intake on cardiometabolic risk markers, gut microbiota and gastrointestinal symptoms in children: A randomized crossover trial. The American Journal of Clinical Nutrition. 2024;119(1):18–28. doi:10.1016/j.ajcnut.2023.10.025

[16] Marshall S, Petocz P, Duve E, Abbott K, Cassettari T, Blumfield M, Fayet-Moore F. The Elect of Replacing Refined Grains with Whole Grains on Cardiovascular Risk Factors: A Systematic Review and Meta-Analysis of Randomized Controlled Trials with GRADE Clinical Recommendation. Journal of the Academy of Nutrition and Dietetics. 2020;120(11):1859–1883.e31. doi:10.1016/j.jand.2020.06.021

[17] Aune D, Keum N, Giovannucci E, Fadnes LT, Boletta P, Greenwood DC, Tonstad S, Vatten LJ, Riboli E, Norat T. Whole grain consumption and risk of cardiovascular disease, cancer, and all cause and cause specific mortality: systematic review and dose-response meta-analysis of prospective studies. BMJ. 2016 Jun 14:i2716. doi:10.1136/bmj.i2716

[18] Aragam KG, Jiang T, Goel A, Kanoni S, Wolford BN, Atri DS, Weeks EM, Wang M, Hindy G, Zhou W, et al. Discovery and systematic characterization of risk variants and genes for coronary artery disease in over a million participants. Nature Genetics. 2022;54(12):1803–1815. doi:10.1038/s41588-022-01233-6

[19] Graham SE, Clarke SL, Wu K-HH, Kanoni S, Zajac GJM, Ramdas S, Surakka I, Ntalla I, Vedantam S, Winkler TW, et al. The power of genetic diversity in genome-wide association studies of lipids. Nature. 2021;600(7890):675–679. doi:10.1038/s41586-021-04064-3

[20] Pazoki R, Vujkovic M, Elliott J, Evangelou E, Gill D, Ghanbari M, Van Der Most PJ, Pinto RC, Wielscher M, Farlik M, et al. Genetic analysis in European ancestry individuals identifies 517 loci associated with liver enzymes. Nature Communications. 2021;12(1):2579. doi:10.1038/s41467-021-22338-2

[21] Ghouse J, Sveinbjörnsson G, Vujkovic M, Seidelin A-S, Gellert-Kristensen H, Ahlberg G, Tragante V, Rand SA, Brancale J, Vilarinho S, et al. Integrative common and rare variant analyses provide insights into the genetic architecture of liver cirrhosis. Nature Genetics. 2024;56(5):827–837. doi:10.1038/s41588-024-01720-y

[22] Yengo L, Sidorenko J, Kemper KE, Zheng Z, Wood AR, Weedon MN, Frayling TM, Hirschhorn J, Yang J, Visscher PM, et al. Meta-analysis of genome-wide association studies for height and body mass index in ∼700000 individuals of European ancestry. Human Molecular Genetics. 2018;27(20):3641– 3649. doi:10.1093/hmg/ddy271

[23] Hartwig FP, Davies NM, Hemani G, Davey Smith G. Two-sample Mendelian randomization: avoiding the downsides of a powerful, widely applicable but potentially fallible technique. International Journal of Epidemiology. 2016;45(6):1717–1726. doi:10.1093/ije/dyx028

[24] Burgess S, Thompson SG. Interpreting findings from Mendelian randomization using the MR-Egger method. European Journal of Epidemiology. 2017;32(5):377–389. doi:10.1007/s10654-017-0255-x

[25] Hemani G, Tilling K, Smith GD. Orienting the causal relationship between imprecisely measured traits using GWAS summary data. PLOS Genetics. 2017;13(11):e1007081. doi:10.1371/journal.pgen.1007081

[26] Morrison J, Knoblauch N, Marcus JH, Stephens M, He X. Mendelian randomization accounting for correlated and uncorrelated pleiotropic elects using genome-wide summary statistics. Nature Genetics. 2020;52(7):740–747. doi:10.1038/s41588-020-0631-4

[27] Darrous L, Hemani G, Davey Smith G, Kutalik Z. PheWAS-based clustering of Mendelian Randomisation instruments reveals distinct mechanism-specific causal elects between obesity and educational attainment. Nature Communications. 2024;15(1):1420. doi:10.1038/s41467-024-45655-8

[28] Shim H, Chasman DI, Smith JD, Mora S, Ridker PM, Nickerson DA, Krauss RM, Stephens M. A Multivariate Genome-Wide Association Analysis of 10 LDL Subfractions, and Their Response to Statin Treatment, in 1868 Caucasians Aspichueta P, editor. PLOS ONE. 2015;10(4):e0120758. doi:10.1371/journal.pone.0120758

[29] GWAS Results. http://www.nealelab.is/uk-biobank/

[30] Results Files for LDSR of UK Biobank GWAS. 2022. https://nealelab.github.io/UKBB_ldsc/downloads.html

[31] Yavorska OO, Burgess S. MendelianRandomization: an R package for performing Mendelian randomization analyses using summarized data. International Journal of Epidemiology. 2017;46(6):1734–1739. doi:10.1093/ije/dyx034

[32] Hemani G, Zheng J, Elsworth B, Wade KH, Haberland V, Baird D, Laurin C, Burgess S, Bowden J, Langdon R, et al. The MR-Base platform supports systematic causal inference across the human phenome. eLife. 2018;7:e34408. doi:10.7554/eLife.34408

[33] Enattah NS, Sahi T, Savilahti E, Terwilliger JD, Peltonen L, Järvelä I. Identification of a variant associated with adult-type hypolactasia. Nature Genetics. 2002;30(2):233–237. doi:10.1038/ng826

[34] Smith CE, Coltell O, Sorlí JV, Estruch R, Martínez-González MÁ, Salas-Salvadó J, Fitó M, Arós F, Dashti HS, Lai CQ, et al. Associations of the MCM6-rs3754686 proxy for milk intake in Mediterranean and American populations with cardiovascular biomarkers, disease and mortality: Mendelian randomization. Scientific Reports. 2016;6(1):33188. doi:10.1038/srep33188

[35] Bjørngaard JH, Nordestgaard AT, Taylor AE, Treur JL, Gabrielsen ME, Munafò MR, Nordestgaard BG, Åsvold BO, Romundstad P, Davey Smith G. Heavier smoking increases colee consumption: findings from a Mendelian randomization analysis. International Journal of Epidemiology. 2017;46(6):1958–1967. doi:10.1093/ije/dyx147

[36] Kamat MA, Blackshaw JA, Young R, Surendran P, Burgess S, Danesh J, Butterworth AS, Staley JR. PhenoScanner V2: an expanded tool for searching human genotype–phenotype associations Kelso J, editor. Bioinformatics. 2019;35(22):4851–4853. doi:10.1093/bioinformatics/btz469

[37] Pirastu N, McDonnell C, Grzeszkowiak EJ, Mounier N, Imamura F, Merino J, Day FR, Zheng J, Taba N, Concas MP, et al. Using genetic variation to disentangle the complex relationship between food intake and health outcomes. Genetics; 2019. http://biorxiv.org/lookup/doi/10.1101/829952. doi:10.1101/829952

[38] Burgess S, Woolf B, Mason AM, Ala-Korpela M, Gill D. Addressing the credibility crisis in Mendelian randomization. BMC Medicine. 2024;22(1):374. doi:10.1186/s12916-024-03607-5

[39] Burgess S, Davey Smith G, Davies NM, Dudbridge F, Gill D, Glymour MM, Hartwig FP, Kutalik Z, Holmes MV, Minelli C, et al. Guidelines for performing Mendelian randomization investigations: update for summer 2023. Wellcome Open Research. 2023;4:186. doi:10.12688/wellcomeopenres.15555.3

[40] Haycock PC, Burgess S, Wade KH, Bowden J, Relton C, Davey Smith G. Best (but oft-forgotten) practices: the design, analysis, and interpretation of Mendelian randomization studies. The American Journal of Clinical Nutrition. 2016;103(4):965–978. doi:10.3945/ajcn.115.118216

[41] Chang M, Yesupriya A, Ned RM, Mueller PW, Dowling NF. Genetic variants associated with fasting blood lipids in the U.S. population: Third National Health and Nutrition Examination Survey. BMC Medical Genetics. 2010;11(1):62. doi:10.1186/1471-2350-11-62

[42] Taba N, Valge H-K, Metspalu A, Esko T, Wilson JF, Fischer K, Pirastu N. Mendelian Randomization Identifies the Potential Causal Impact of Dietary Patterns on Circulating Blood Metabolites. Frontiers in Genetics. 2021 [accessed 2022 Nov 22];12. https://www.frontiersin.org/articles/10.3389/fgene.2021.738265

[43] Mohanty SP, Singhal G, Scuccimarra EA, Kebaili D, Héritier H, Boulanger V, Salathé M. The Food Recognition Benchmark: Using Deep Learning to Recognize Food in Images. Frontiers in Nutrition. 2022;9:875143. doi:10.3389/fnut.2022.875143

[44] Karabay A, Varol HA, Chan MY. Improved food image recognition by leveraging deep learning and data-driven methods with an application to Central Asian Food Scene. Scientific Reports. 2025;15(1):14043. doi:10.1038/s41598-025-95770-9

[45] Konstantakopoulos FS, Georga EI, Fotiadis DI. A novel approach to estimate the weight of food items based on features extracted from an image using boosting algorithms. Scientific Reports. 2023;13(1):21040. doi:10.1038/s41598-023-47885-0

