## Supplementary material for "Dietary Intake Mendelian Randomization: Assessment and Development of Methods for Instrument Selection and Robust Inference": STROBE-MR Checklist

**STROBE-MR checklist of recommended items to address in reports of Mendelian randomization studies**[1 2]

| Item No. | Section | Checklist item | Page No. | Relevant text from manuscript |
|---|---|---|---|---|
| 1 | **TITLE and ABSTRACT** | Indicate Mendelian randomization (MR) as the study's design in the title and/or the abstract if that is a main purpose of the study | 1-3 | **Title:** Dietary Intake Mendelian Randomization: Assessment and Development of Methods for Instrument Selection and Robust Inference<br><br>**Abstract:**<br>**Background:** Mendelian randomization (MR) uses genetic instruments (GI) to infer causality between exposures, like dietary intake, and health outcomes. Almost all MR of dietary intake use the full set of genome-wide significant (GWS) variants in the GI, and therefore, causal estimates are likely biased by variants that act indirectly on diet.<br><br>**Objective:** First, we performed an assessment of the diet MR literature to evaluate the applications and approaches common in the field. Second, using conventional two-sample MR techniques with GWS variants, we evaluated whether MR could detect expected associations between six diet-health relationships supported by existing nutrition science literature. Third, we developed and tested methods for refining the GI using filtering and mediation-based approaches.<br><br>**Methods:** Studies that performed MR of foods or beverages on any health outcome were identified in PubMed. We recorded how the GI was created, what dietary intake traits were studied, how the exclusion restriction assumption was evaluated, and what sensitivity tests were performed. We tested if conventional MR methods could detect established diet-health relationships by selecting a biomarker and disease outcome for each dietary trait (six positive controls total). This included oily fish intake on triglycerides (TG) and cardiovascular disease (CVD), alcohol intake on alanine aminotransferase (ALT) and liver cirrhosis, and white vs whole grain or brown bread on LDL cholesterol (LDL-C) and CVD. To refine the GI to better estimate the direct effect of diet by removing or accounting for the indirect effects of confounders, we tested two phenome-wide association study (PheWAS) based GI filtering approaches and a mediation approach via multivariable MR (MVMR). Causal inferences were estimated by the inverse variance weighted (IVW) and weighted median (WM) estimators and by MR-CAUSE.<br><br>**Results:** There is a strong and rapidly expanding interest in applying MR to dietary intake exposures (178 studies identified with 76 published in 2024). Existing studies showed a wide range of methodological rigor, especially with respect to GI specificity, which raised concerns whether MR using GWS GIs can adequately evaluate diet-health relationships. In empirical testing, conventional two-sample MR methods on GWS GIs only identified the relationships between oily fish on TG and white vs whole grain or brown bread on LDL-C using the WM estimator, whereas no relationships were identified by the IVW estimator. Filtering the GI improved the ability to detect the expectation for diet-biomarker pairs (IVW, oily fish on TG: ß=-0.12 [95% CI -0.18 to -0.054]; IVW, white vs whole grain or brown bread on LDL-C: ß = 0.11 [95% CI 0.058 to 0.16]) but not diet-disease pairs. MR- |

| | | | | |
|---|---|---|---|---|
| | | | | CAUSE identified the only diet-disease association – white vs. whole grain or brown bread on CVD ($\gamma$=0.17 [95% credible interval, 0.09 to 0.25]). Furthermore, MR-CAUSE found that many diet-health relationships were impacted by confounding. We evaluated which traits contributed to confounding via the PheWAS results and found that body composition traits were the most prevalent confounders. The PheWAS output was used to prioritize traits for MVMR and rescued the expected direct effect of alcohol on ALT (ß= 0.028 [95% CI 0.017 to 0.039]).<br><br>**Conclusion:** MR studies of diet's causal role in health have flooded the literature; however, our inconsistent associations with positive and negative controls using multiple tests and filtering methods signal a need for caution. More thoughtful curation of the GI is critical to reduce confounding due to health and environmental factors when evaluating the causal effect of diet on health. |
| | **INTRODUCTION** | | | |
| 2 | **Background** | Explain the scientific background and rationale for the reported study. What is the exposure? Is a potential causal relationship between exposure and outcome plausible? Justify why MR is a helpful method to address the study question | 4 | Background and rationale: Genetic instruments (GIs) for dietary intake traits are publicly available and MR of these traits on health outcomes is increasingly common. However, dietary intake is affected by both genetics and environmental factors, and we suspect that many of the variants in the GI affect diet through indirect paths (a violation of the exclusion-restriction assumption).<br><br>Exposure: The exposures included oily fish intake (ordinal, described by frequency from never to daily intake), bread type most consumed (ordinal, described by food types), and alcohol intake (continuous, described by never to daily intake). The exposures are described in detail in Supplemental Table 1.<br><br>Plausibility: Diet has profound effects on health, but many diet-health relationships are unsettled. With the availability of GI for dietary intake traits (food groups, foods, food liking, etc.) dietary intake MR are becoming more common. However, dietary intake is influenced by non-genetic factors such as socioeconomic status and health status. We hypothesize that many of the variants statistically associated with diet act through indirect paths and violate the exclusion-restriction assumption. |
| 3 | **Objectives** | State specific objectives clearly, including pre-specified causal hypotheses (if any). State that MR is a method that, under specific assumptions, intends to estimate causal effects | 4 | The objectives of our study are included in the abstract and in the introduction.<br><br>First, we performed an assessment of the diet MR literature to evaluate the applications and approaches common in the field. Second, using conventional two-sample MR techniques with GWS variants, we evaluated whether MR could detect expected associations between six diet-health relationships supported by existing nutrition science literature. Third, we developed and tested methods for refining the GI using filtering and mediation-based approaches. |

| | | | | |
|---|---|---|---|---|
| | | | | We have stated that MR is a method that intends to estimate causal effects given specific statistical assumptions are met in the introduction. |
| | **METHODS** | | | |
| 4 | **Study design and data sources** | Present key elements of the study design early in the article. Consider including a table listing sources of data for all phases of the study. For each data source contributing to the analysis, describe the following: | 5-8 | Figure 1 provides an overview of the study design. Supplemental Table 1 (ST1) includes the data sources for all phases of the study. ST1 includes relevant information about the genome-wide association study (GWAS) summary data such as the transformation of the original variable, the units, the covariates from the GWAS, whether the variable was ordinal, continuous, or binary, and the cohort the data was derived from. |
| | a) | Setting: Describe the study design and the underlying population, if possible. Describe the setting, locations, and relevant dates, including periods of recruitment, exposure, follow-up, and data collection, when available. | 5-6 | Study design: The design of the study was 3-fold. First, to evaluate the dietary intake MR literature; second to test the ability of genome-wide significant (GWS) GIs for dietary intake to detect nutrition science-supported relationships between dietary intake and health; and third, to refine the GI to identify the direct effect of dietary intake on health using GI filtering and mediation approaches.<br>Exposure: The summary statistics for the exposure GWAS were obtained from a study completed using the UK Biobank (Cole, Nature Comm, 2020).<br>Outcome: The summary statistics for the outcome GWASs were obtained from the GWAS catalog and are described in ST1. |
| | b) | Participants: Give the eligibility criteria, and the sources and methods of selection of participants. Report the sample size, and whether any power or sample size calculations were carried out prior to the main analysis | 6-7 | Summary statistic data was used from publicly available sources. Thus, decisions about eligibility criteria were not completed for the present study. We did assess the covariates, transformations, and eligibility requirements from the published GWAS to ensure we selected data with minimum risk of collider bias. |
| | c) | Describe measurement, quality control and selection of genetic variants | 6-7 | In objective 2, we used independent GWS GIs prioritized from the exposure GWAS. In objective 3, we began with the GWS GI and then filtered based on results from the PheWAS-PVE, PheWAS-Ttest, and the Core test (described in the methods). We tested the methods on dietary intake-health relationships supported by nutrition science as an indicator of performance. |
| | d) | For each exposure, outcome, and other relevant variables, describe methods of assessment and diagnostic criteria for diseases | | This is provided in Supplemental Table 1. |

| | | | | | |
|---|---|---|---|---|---|
| | | e) | Provide details of ethics committee approval and participant informed consent, if relevant | | NA |
| 5 | **Assumptions** | | Explicitly state the three core IV assumptions for the main analysis (relevance, independence and exclusion restriction) as well assumptions for any additional or sensitivity analysis | 4 | The three core IV assumptions are stated in the introduction. |
| 6 | **Statistical methods: main analysis** | | Describe statistical methods and statistics used | | |
| | | a) | Describe how quantitative variables were handled in the analyses (i.e., scale, units, model) | | Descriptions of how each exposure and outcome variable were processed is provided in Supplemental Table 1. |
| | | b) | Describe how genetic variants were handled in the analyses and, if applicable, how their weights were selected | 6 | GWAS exposure and outcome data was quality checked and harmonized using custom scripts. Effect alleles for the dietary intake and outcome GWAS data were flipped to the same direction as needed. Ambiguous variants were removed if their minor allele frequency was between 0.45 to 0.55. Proxy variants in linkage disequilibrium (r2>0.8) with the original dietary intake variant were used when the original variant was missing in the outcome GWAS. In these cases, the dietary intake variant was also replaced with the proxy to avoid problems of phasing. |
| | | c) | Describe the MR estimator (e.g. two-stage least squares, Wald ratio) and related statistics. Detail the included covariates and, in case of two-sample MR, whether the same covariate set was used for adjustment in the two samples | 6 | We employed the inverse variance weighted and the weighted median methods, which use two-stage least squares. We also employed MR-CAUSE. |
| | | d) | Explain how missing data were addressed | 6 | When a variant from the GI was missing in the outcome data, we attempted to find a proxy using the following strategy.<br>Proxy variants in linkage disequilibrium ($r^2$>0.8) with the original exposure variant were identified for variants that were missing in the outcome GWAS. In these cases, the exposure variant was also replaced with the proxy to avoid problems of phasing. The R package LDLinkR was used with the Great Britain reference population in build 37. When multiple proxies with identical $r^2$ values were |

| | | | | |
|---|---|---|---|---|
| | | | | available, the variant closest to the original was selected. If the variant was not in the 1000G reference panel, it was removed from the GI. |
| | | e) If applicable, indicate how multiple testing was addressed | 6 | We evaluated 9 relationships. Thus, we employed a multiple testing correction of P<0.05/9=0.0055. This is a conservative threshold as the biomarkers and diseases are related. |
| 7 | **Assessment of assumptions** | Describe any methods or prior knowledge used to assess the assumptions or justify their validity | 6-8 | The third aim of our study was designed to address the exclusion restriction assumption. We tested various GI filtering methods using external PheWAS data and used multivariable MR. |
| 8 | **Sensitivity analyses and additional analyses** | Describe any sensitivity analyses or additional analyses performed (e.g. comparison of effect estimates from different approaches, independent replication, bias analytic techniques, validation of instruments, simulations) | 6-8 | We evaluated pleiotropy using the MR-Egger intercept test (Supplemental Table 4) and MR-CAUSE (Table 4 and 5). We calculated heterogeneity via Cochran's Q statistic and evaluated the F statistic. |
| 9 | **Software and pre-registration** | | | |
| | | a) Name statistical software and package(s), including version and settings used | 8 | The statistical software and packages used are described in the methods including specific settings for the functions within the R packages MendelianRandomization and MRCAUSE.<br>RStudio (v 4.2.2)<br>R packages: MendelianRandomization (v 0.10.0), MR-CAUSE (v 1.2.0)<br>IVW: random effects and default setting<br>MR-CAUSE: the nuisance parameters were calculated using 1,000,000 variants without replacement. The genome-wide variants were pruned with the ld_prune function with the 1000 Genome CEU population and a statistical threshold of 5E-3. |
| | | b) State whether the study protocol and details were pre-registered (as well as when and where) | | NA |
| | **RESULTS** | | | |
| 10 | **Descriptive data** | | | |
| | | a) Report the numbers of individuals at each stage of included studies | | NA |

| | | | | | |
|---|---|---|---|---|---|
| | | | and reasons for exclusion. Consider use of a flow diagram | | |
| | | b) | Report summary statistics for phenotypic exposure(s), outcome(s), and other relevant variables (e.g. means, SDs, proportions) | | See Supplemental Table 1. |
| | | c) | If the data sources include meta-analyses of previous studies, provide the assessments of heterogeneity across these studies | | NA |
| | | d) | For two-sample MR:<br>i. Provide justification of the similarity of the genetic variant-exposure associations between the exposure and outcome samples<br>ii. Provide information on the number of individuals who overlap between the exposure and outcome studies | | The exposure and outcome data including the cohorts they were derived from are described in Supplemental Table 1.<br>The exposure data is from the UK Biobank. Of the six outcomes analyzed, only one (alanine aminotransferase) is composed solely of the UK Biobank. This study validated their GWAS using data obtained from the Rotterdam, Lifelines, and Million Veterans Program. This ALT GWAS was selected despite the overlap due to its size, availability, and recent publication date.<br>Overlap between the diet and triglyceride and LDL cholesterol GWAS is estimated at ~17%.<br>It is difficult to estimate the overlap between the diet and cardiovascular disease, liver cirrhosis, and height GWAS as each were meta-analyses consisting of numerous cohorts including the UKB. |
| 11 | **Main results** | | | | |
| | | a) | Report the associations between genetic variant and exposure, and between genetic variant and outcome, preferably on an interpretable scale | 10-11 | All of the GIs are available in Supplemental Table 2. |
| | | b) | Report MR estimates of the relationship between exposure and outcome, and the measures of uncertainty from the MR analysis, on an interpretable scale, such as odds ratio or relative risk per SD difference | | We originally sought to compare the effects of the MR and RCT studies. However, the ordinal nature of the diet traits, coupled with the difficulty of identifying RCTs solely testing the food of interest, made this goal unachievable. |

| | | | | | |
|---|---|---|---|---|---|
| | | c) | If relevant, consider translating estimates of relative risk into absolute risk for a meaningful time period | | Not relevant. |
| | | d) | Consider plots to visualize results (e.g. forest plot, scatterplot of associations between genetic variants and outcome versus between genetic variants and exposure) | | Forrest plots for all trait pairs are included in Figures 3 and 4. |
| 12 | **Assessment of assumptions** | | | | |
| | | a) | Report the assessment of the validity of the assumptions | | The assessment of the exclusion restriction assumption is discussed in the discussion. |
| | | b) | Report any additional statistics (e.g., assessments of heterogeneity across genetic variants, such as $I^2$, Q statistic or E-value) | 10-12 | Heterogeneity via Cochrans Q statistic is reported from the IVW tests on the GWS and filtered GI in Table 2. MR-Egger intercept tests are provided in Supplemental Table 4. |
| 13 | **Sensitivity analyses and additional analyses** | | | | |
| | | a) | Report any sensitivity analyses to assess the robustness of the main results to violations of the assumptions | 10-12 | Aim 3 evaluated whether filtering the GI based on external PheWAS data or adjusting for the indirect effects of confounding traits could improve the detection of expected nutrition science relationships. |
| | | b) | Report results from other sensitivity analyses or additional analyses | 10-12 | The results from the filtered GI are provided in Tables 2 and 3, Figures 3 and 4, Table 7, and Supplemental Table 9. |
| | | c) | Report any assessment of direction of causal relationship (e.g., bidirectional MR) | | We did not conduct bidirectional MR. We did employ Steiger filtering, which addresses the likely main direction a variant within the GI is acting (X→Y or Y→X). |

| | | | | |
|---|---|---|---|---|
| | | d) When relevant, report and compare with estimates from non-MR analyses | | We compared the direction of the relationship with the existing literature. See Table 1, 2, 3 and the discussion. |
| | | e) Consider additional plots to visualize results (e.g., leave-one-out analyses) | | |
| | **DISCUSSION** | | | |
| 14 | **Key results** | Summarize key results with reference to study objectives | | |
| 15 | **Limitations** | Discuss limitations of the study, taking into account the validity of the IV assumptions, other sources of potential bias, and imprecision. Discuss both direction and magnitude of any potential bias and any efforts to address them | 15 | Limitations of the study are included in the discussion. Our study was conducted in individuals of European genetic similarity. The transferability of the results is unclear due to both genetic and dietary factors being tied to genetic and cultural factors, respectively. Further, we used nutrition science to steer the positive and negative controls we selected but needed the relationships prioritized by the literature to have publicly available summary statistics from a recently conducted GWAS in a large, European, non-UKB cohort. We originally sought to compare the effects of the MR and RCT studies. However, the ordinal nature of the diet traits, coupled with the difficulty of identifying RCTs solely testing the food of interest, made this goal unachievable. The IVW test requires the no measurement error (NOME) assumption, which is intrinsically difficult when measuring dietary intake by recall. |
| 16 | **Interpretation** | | | |
| | | a) Meaning: Give a cautious overall interpretation of results in the context of their limitations and in comparison with other studies | 16 | Concern about the credibility of diet-MR is timely due to the growing interest in this field. Due to the inconsistency and inaccuracy of the positive and negative controls across multiple MR estimators, MR using GWS GIs derived from food frequency questionnaires or 24-hour dietary assessments should be used and interpreted with caution. Scrutiny of the GI is paramount to avoid mischaracterizing the causal relationships between dietary intake and health and further eroding public trust in nutrition science. |
| | | b) Mechanism: Discuss underlying biological mechanisms that could drive a potential causal relationship between the investigated exposure and the outcome, and whether the gene-environment equivalence assumption is reasonable. Use causal language carefully, | 5 | Oily fish on TG and CVD is via omega-3. White vs whole grain or brown bread on LDL-C and CVD is via fiber. Alcohol on ALT and liver cirrhosis is via ethanol. |

| | | | | |
|---|---|---|---|---|
| | | clarifying that IV estimates may provide causal effects only under certain assumptions | | |
| | | c) Clinical relevance: Discuss whether the results have clinical or public policy relevance, and to what extent they inform effect sizes of possible interventions | 16 | Due to the inconsistency and inaccuracy of the positive and negative controls across multiple MR estimators, MR using GWS GIs derived from food frequency questionnaires or 24-hour dietary assessments should be used and interpreted with caution. Scrutiny of the GI is paramount to avoid mischaracterizing the causal relationships between dietary intake and health and further eroding public trust in nutrition science. |
| 17 | **Generalizability** | Discuss the generalizability of the study results (a) to other populations, (b) across other exposure periods/timings, and (c) across other levels of exposure | 15 | Due to linkage disequilibrium, minor allele frequency, and data availability, these analyses were restricted to GWAS conducted in individuals with European genetic ancestry. Additionally, culturally appropriate dietary questionnaires in populations with different ancestry backgrounds would likely result in the use of a different FFQ, making the harmonization of phenotypes for future meta-analyses difficult. |
| | **OTHER INFORMATION** | | | |
| 18 | **Funding** | Describe sources of funding and the role of funders in the present study and, if applicable, sources of funding for the databases and original study or studies on which the present study is based | 17 | Research reported in this publication was supported by the National Library Of Medicine and the National Institute of Diabetes and Digestive and Kidney Diseases of the National Institutes of Health under Award Numbers T15LM009451, T32DK007658-35, and R00DK127196. The content is solely the responsibility of the authors and does not necessarily represent the official views of the National Institutes of Health. |
| 19 | **Data and data sharing** | Provide the data used to perform all analyses or report where and how the data can be accessed, and reference these sources in the article. Provide the statistical code needed to reproduce the results in the article, or report whether the code is publicly accessible and if so, where | | Data described in the manuscript are publicly and freely available without restriction at the GWAS catalog (Supplemental Table 1: LDL-C, GCST90239658; CVD, GCST90132314; TG, GCST90239664; ALT, GCST90013405; Cirrhosis of liver, GCST90013405; Height, GCST006901) and the Type 2 Diabetes Knowledge Portal by the dataset name, UK Biobank dietary habit GWAS. |
| 20 | **Conflicts of Interest** | All authors should declare all potential conflicts of interest | | No authors have conflicts of interest. |

This checklist is copyrighted by the Equator Network under the Creative Commons Attribution 3.0 Unported (CC BY 3.0) license.

1. Skrivankova VW, Richmond RC, Woolf BAR, Yarmolinsky J, Davies NM, Swanson SA, et al. Strengthening the Reporting of Observational Studies in Epidemiology

using Mendelian Randomization (STROBE-MR) Statement. JAMA. 2021;under review.

2. Skrivankova VW, Richmond RC, Woolf BAR, Davies NM, Swanson SA, VanderWeele TJ, et al. Strengthening the Reporting of Observational Studies in Epidemiology using Mendelian Randomisation (STROBE-MR): Explanation and Elaboration. BMJ. 2021;375:n2233.
